# Differential effects of sodium channel blockers on *SCN8A* gain-of-function variants associated with drug-responsive or -resistant epilepsy

**DOI:** 10.64898/2026.05.21.26353295

**Authors:** Hang Lyu, Siyu Li, Roberto Previtali, Katrine M. Johannesen, Bingqi Guo, Chrisitian M. Boßelmann, Elena Gardella, Rikke S Møller, Holger Lerche, Yuanyuan Liu

## Abstract

Gain-of-function variants (GOF) in *SCN8A*, which encodes the Na_V_1.6 sodium channel, lead to epilepsy syndromes ranging from drug-responsive self-limited (SeLIE) and intermediate epilepsy to drug-resistant developmental and epileptic encephalopathy (DEE). It is currently unclear why individuals with *SCN8A* GOF variants show variable responses to sodium channel blockers (SCBs). Here, we compared the clinical characteristics of 173 individuals with 25 different *SCN8A* GOF variants following the hypothesis that carriers of variants affecting activation gating respond less well to SCBs than those with variants affecting fast inactivation gating, given that use-dependent SCBs preferentially target inactivated channel states. We found that individuals with variants altering channel activation gating were more severely affected than those with variants altering inactivation properties: They had an earlier age at onset (3 vs. 5 months, *P* < 0.0001), higher prevalence of DEE (75% vs. 39%; *P* < 0.0001), and poorer response to SCBs (20% vs. 69% seizure free; *P* < 0.0001). We performed pharmacological studies on representative and recurrent variants from each group: two variants (F846S and M1760I) causing hyperpolarizing shifts of the voltage-dependent activation curves, and two variants (G1475R and N1877S) causing depolarizing shifts of the voltage-dependent fast inactivation curves. Phenytoin failed to suppress neuronal firing in neurons expressing activation-related variants, but showed good suppressing effects in neurons expressing inactivation-related variants. In contrast, PRAX-330, a new SCB, which showed much faster binding rates than phenytoin, was effective for both groups of variants by markedly reducing neuronal firing through rapidly and persistently stabilizing Na_V_1.6 in the inactivated state. Our findings provide new insights into the mechanism of drug-resistance in *SCN8A*-DEE and support PRAX-330 and compounds with similar pharmacological properties as a promising preclinical candidate for targeted therapies.

## Introduction

Gain-of-function (GOF) variants in *SCN8A*, which encodes the Na_V_1.6 sodium channel, have been implicated in a wide spectrum of epileptic syndromes, with the severity of clinical manifestations exhibiting a clear correlation to functional changes at biophysical level^1,2^. *SCN8A* GOF variants causing mild changes of activation or inactivation properties typically result in self-limited (familial) infantile epilepsy (SeLIE; previously benign familial infantile epilepsy, BFIE) or epilepsies of intermediate severity^3–6^; meanwhile, GOF variants causing strong alterations of activation and inactivation properties in combination with other biophysical changes (e.g. increase of persistent current, acceleration of recovery) are associated with intermediate epilepsy or severe developmental and epileptic encephalopathy (DEE)^7–10^. Using a single-compartment conductance-based model to simulate neuronal firing, we found that changes of activation properties have a stronger effect than the changes of inactivation properties on neuronal firing^1^. The electrophysiological scores determined by biophysical changes of *SCN8A* GOF variants are significantly higher for the patients with DEE than the patients with mild or intermediate epilepsy^1,2^.

Interestingly, not all affected individuals carrying *SCN8A* GOF variants respond to sodium channel blockers (SCBs). In fact, quite many individuals with *SCN8A* GOF variants still have drug-resistant epilepsy^11^, i.e. they continue to have seizures despite >2 trials of two or more tolerated and appropriately chosen anti-seizure medications (ASMs), whereas others exhibit favorable treatment responses. For example, two individuals harboring GOF variants that primarily induce a hyperpolarizing shift of the activation curve, c.5280G>A (p.Met1760Ile, M1760I) and c.2537T>C (p.Phe846Ser, F846S), presented with early-onset, refractory seizures and suffered from severe developmental delay and premature death despite early treatment with phenytoin (PHT), carbamazepine (CBZ) and other ASMs^6,12^. In contrast, two individuals carrying GOF variants that cause a depolarizing shift of the fast inactivation curve, c.4423G>A (p.Gly1475Arg, G1475R) and c.5630A>G (p.Asn1877Ser, N1877S), developed focal epilepsy with onset before 1 year of age, and achieved seizure freedom with monotherapy using oxcarbazepine (OXC) or lamotrigine (LTG)^6,7^. These divergent therapeutic outcomes and the known mechanism of action of use-dependent SCBs binding to and stabilizing the fast inactivated state of voltage-gated sodium (Na_V_) channels, raised the hypothesis that the distinct biophysical properties of *SCN8A* GOF variants, particularly their differential effects on activation and inactivation gating, underlie the variability in drug-responses.

Biophysically, classical SCBs such as PHT, CBZ, OXC, and LTG, inhibit Na_V_ channels by preferentially binding to their fast inactivated state in a use- and voltage-dependent manner. This mechanism selectively suppresses high-frequency repetitive firing while preserving normal neuronal activity^13–17^. Nevertheless, classical SCBs exhibit limited therapeutic efficacy in individuals with DEE, and long-term or high-dose administration is often associated with significant safety concerns. A new inhibitor of Na_V_ channels, PRAX-330 (known before as GS967), has been shown to attenuate the enhanced persistent current and abnormal neuronal firing in two *Scn8a*-DEE mouse models, resulting in reduced seizure frequency and prolonged survival^18–20^. These findings highlight its therapeutic potential for severe epileptic encephalopathies. However, despite these promising preclinical outcomes, the biophysical and neuronal mechanisms underlying the efficacy of PRAX-330 compared to classical SCBs remain insufficiently understood.

In the present work, we collected clinical phenotypes and therapeutic outcomes of individuals with *SCN8A* GOF variants from our own and further previously published work^1,6,9,10,21–25^ to explore the correlation of the drug-responses with the biophysical changes of the GOF variants (activation- vs. inactivation-alterations). Using PHT as a representative of classical SCBs, we further evaluated the biophysical properties and neuronal firing behavior for Na_V_1.6 channels harboring the variant F846S or M1760I (activation-group) versus G1475R or N1877S (inactivation-group) upon the administration of PHT versus PRAX-330 respectively. Our results provide a new insight into the mechanisms of drug-resistant epilepsy caused by *SCN8A* GOF variants and suggest a much stronger in-vitro efficacy of PRAX-330 compared with PHT, regardless of the GOF mechanism.

## Materials and methods

### Clinical cohort phenotyping

Individuals with *SCN8A* variants included in this study were collected from previously published cases as well as from our characterized cohort^1^. For the published cases, we searched the PubMed database using the keyword “SCN8A” and included all studies reporting original cases or probands (summarized in Supplementary Table 1). To avoid duplicate inclusion of affected individuals, we contacted referring physicians and/or corresponding authors when necessary. The latest search was performed on July 1, 2025. Individuals from our own cohort were recruited through a collaborative clinical network, and their clinical characteristics, EEG, neuroimaging and treatment data were retrospectively assessed using a standardized phenotyping sheet^1^. Seizures and epilepsy syndromes were classified according to the latest ILAE guidelines^26,27^, with the understanding that data from previous studies often preceded the guidelines. Thus, information on onset and awareness was not consistently available.

Functional data of GOF variants included in this study were derived from previously published reports as well as our unpublished electrophysiological results. The affected individuals were categorized into two groups based on the biophysical changes of their respective GOF variants: (i) Activation group, comprising individuals carrying variants that induce a hyperpolarizing shift of the steady-state activation curve along with additional biophysical alterations; (ii) Inactivation group, comprising individuals carrying variants that cause a depolarizing shift of the steady-state inactivation curve in combination of other biophysical changes. There are three GOF variants (p.(Ile1327Val), p.(Arg1617Gln) and p.(Arg1872Gln)) that cause a hyperpolarizing shift of the activation curve as well as a depolarizing shift of the inactivation curve. Considering that the alterations of activation properties typically exert a more dominant influence on overall channel function based on the model results in our previous study^6^, individuals with these variants were classified into the activation group.

Treatment response was evaluated by the referring treatment providers and grouped into four categories: 1. seizure free for at least 6 months following treatment); 2. seizure reduction with worthwhile clinical improvement noted by clinicians and caregivers, treated individuals stayed on the drug (a structured information on >50% responder rate was not available); 3. no effect, i.e. treatment discontinued due to lack of benefit; 4. worsening with aggravation of seizures (reported by providers and caregivers). Among all recorded ASMs, phenytoin, oxcarbazepine, carbamazepine, lamotrigine, zonisamide, and lacosamide were classified as SCBs, whereas levetiracetam, valproate, topiramate, clobazam, phenobarbital, vigabatrin, clonazepam, and ketogenic diet therapy were categorized as non-SCBs.

### Ethics

The study was approved by the local ethical committees. Animal protocols for the primary neuronal cultures were approved by the local Animal Care and Use Committee (Regierungspraesidium Tuebingen)

### In vitro pharmacological studies

Based on the clinical phenotypes of the affected individuals, the recurrence frequency of the variants and the functional alterations, we selected two variants (F846S, M1760I) from the activation group and two variants (G1475R, N1877S) from the inactivation group for pharmacological investigations. The constructs of wild-type (WT) and the selected variants, which are tetrodotoxin resistant, had been generated in our previous studies^6,17^. Either WT or mutant Na_V_1.6 channels were transfected into neuroblastoma (ND7/23) cells together with the β1- and β2-subunits using Lipofectamine 2000 (Invitrogen); or into murine cultured hippocampal neurons together with GFP under the promoter targeting excitatory neurons (pAAV-CAMKII-GFP, plasmid #6454; Addgene) using Optifect (Invitrogen)^28^. Whole-cell patch clamp recordings were performed to obtain the biophysical parameters and neuronal properties for WT and mutant Na_V_1.6 channels as previously described^6,17^. The detailed protocols are summarized in Supplementary Materials and Methods.

Phenytoin (PHT, Sigma-Aldrich) and PRAX-330 (MedChemExpress) were dissolved in dimethyl sulfoxide (DMSO) respectively and stored in aliquots at −20. Prior to use, stock solutions were diluted to target concentration with electrophysiological bath solutions. Bath solutions containing only DMSO were used for the control solutions. The final concentration of DMSO in the control and drug-containing bath solutions was kept at 0.1% to avoid the toxic effect of DMSO and make all experiments comparable. Unpaired recordings were performed for most pharmacological experiments except for drug binding and unbinding experiments in ND7/23 cells.

For the latter, cells were first stabilized in bath solutions without DMSO for 5 min, then perfused with either control or drug-containing bath solution using a manual syringe-driven perfusion system. A minimum equilibration time of 5 min was required before the voltage-step protocols (Supplementary Fig. 1A and E) were applied to the cells. Each cell was tested with only one drug concentration. Cells exhibiting significant current rundown (>25%) or unstable series resistance after drug application were excluded from analysis.

### Data recording and statistical analysis

Clinical data were analyzed using Microsoft Excel (Microsoft Corporation) and GraphPad Prism 8.0 (GraphPad Software). χ^2^ tests (pie chart) or Fisher’s exact test (bar chart) were used for categorical data. Comparison between two groups of ordinal variables were assessed using the Mann-Whitney U test.

Electrophysiological data were analyzed with Clampfit software of pClamp 10.6 (Axon Instruments), Microsoft Excel and Graphpad prism 8.0. The D’Agostino & Pearson test was used to assess normality assumption. For statistical evaluation of two groups, an independent t-test was applied for normally distributed and a Mann-Whitney U test for not normally distributed data. For comparison of normally distributed multiple groups, an ordinary one-way or two-way ANOVA with post hoc test was used for the data with equal SDs, while Welch’s correction was applied when data exhibited unequal SDs; For comparison of not normally distributed multiple groups, a Kruskal-Wallis with Dunn’s post hoc test was applied. For all groups with significance compared to controls, exact P values were reported; any P values less than 0.0001 were reported as “*P* < 0.0001”.

## Results

### Clinical phenotypes and responsiveness to ASMs in individuals with *SCN8A* GOF variants

The study cohort consisted of 173 individuals, each carrying one of 25 *SCN8A* GOF variants which have been previously functionally characterized. In total, 16 activation-related *SCN8A* GOF variants (106 individuals), and nine inactivation-related *SCN8A* GOF variants (67 individuals) were counted. These activation-related (yellow) and inactivation-related (blue) GOF variants are distributed across the entire *SCN8A* structure (Fig. 1A). The variants in the activation group caused a hyperpolarizing shift of the steady-state activation curve by 2.5 to 11.3 mV, whereas those in the inactivation group induced a depolarizing shift of the steady-state inactivation curve by 3.8 to 13.4 mV. A summary of clinical phenotypes and pharmacological outcomes is shown in Fig. 1B-H. Detailed clinical data is provided in Supplementary Table 1.

**Fig. 1:**
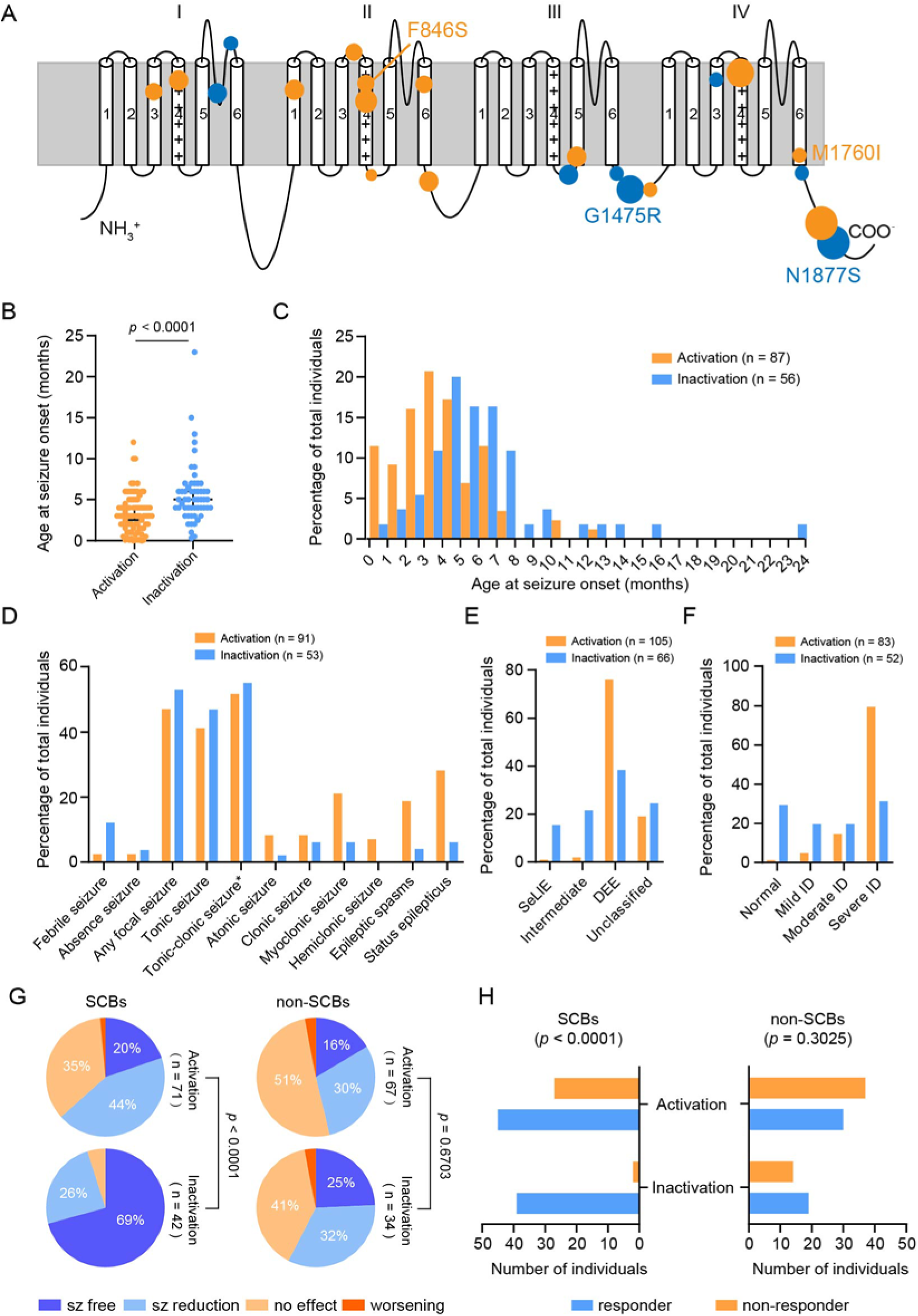
Location of *SCN8A* GOF variants included in this study and clinical phenotypes of individuals with these variants. (A) Schematic 2D representation of the Na_V_1.6 channel displaying the location of activation-related GOF variants (yellow) and inactivation-related GOF variants (blue). Recurrent variants are depicted with symbol sizes proportional to the number of affected individuals. The four selected variants (M1760I, F846S, G1475R, N1877S) for further functional and pharmacological studies are specifically indicated. (B) Scatter plot of the age at seizure onset of affected individuals from activation versus inactivation group. (C) Histogram illustrating the different distributions of age at seizure onset between the two groups. Histogram bin size = 1 month. The activation and inactivation groups also showed different distribution of seizure types (D), epileptic phenotypes (E) and intellectual disability (ID) severity levels. (G) and (H) Treatment responses to SCBs and non-SCBs in affected individuals with *SCN8A* GOF variants. P values derived from χ2 test are provided under the pie chart (G) or from Fisheŕs exact test are provided above the bar chart (H). Tonic-clonic seizure*: Tonic-clonic seizures with unknown onset.

For the activation group, the median age at seizure onset was three months. Nine (8.5%) individuals were prematurely deceased at the median age of 22 months. Eighty (75%) individuals were diagnosed with DEE, while 23 (22%) had epilepsy not further classified. SeLIE (1%) and intermediate epilepsy (2%) were rare. The most frequent seizure type was tonic-clonic seizures (52%) (Supplementary Table 1). Cognition ranged from normal (1%), mild/ moderate intellectual disability (ID) (19%) to severe ID (80%). SCBs response was reported for 71 individuals with 14 (20%) achieving seizure freedom, 31 (44%) seizure reduction, and 25 (35%) with no effect. Seizure exacerbation was reported in one (1%) individual. Non-SCBs response was reported for 67 individuals, with 11 (16%) achieving seizure freedom, 20 (30%) seizure reduction, 34 (51%) had no effect, and two (3%) reporting seizure exacerbation. Most variants occurred *de novo* (88%), while inheritance was unknown in 12 individuals. Fourteen variants were recurrent: p.(Arg1872Trp/Gln/Leu) (n=44) and p.(Arg1617Gln) (n=17) were the most common variants.

For the inactivation group, the median age at seizure onset was five months. One individual died in early childhood, another individual died during adolescence. Twenty-six (39%) individuals had DEE, 16 (24%) epilepsy not further classified, 14 (21%) intermediate epilepsy, 10 (15%) SeLIE, and only one individual did not have epilepsy but showed severe ID. The most frequent seizure type was tonic-clonic seizures (55%) (Supplementary Table 1). Cognition ranged from normal (29%), mild/ moderate ID (40%) to severe ID (31%). SCBs effect was reported for 42 individuals, with 29 (69%) achieving seizure freedom, 11 (26%) seizure reduction and two (5%) had no response. Non-SCBs effect was reported for 34 individuals, with eight individuals (25%) achieving seizure freedom, 11 (32%) seizure reduction, 14 (32%) had no response, and one (2%) report seizure exacerbation. Most variants occurred *de novo* (66%), while seven individuals were familial cases. Inheritance was unknown in thirteen individuals. Five variants were recurrent: p.(Gly1475Arg) (n=24) and p.(Asn1877Ser) (n=28) were the most common variants (Fig. 1A).

The activation group exhibited significantly earlier seizure onset than the inactivation group (Fig. 1B, Mann-Whitney test, *P* < 0.0001). The histograms of the age at seizure onset also show a clear bimodal distribution for these two groups (Fig. 1C). Febrile seizures were more frequent in the inactivation group, whereas myoclonic seizures, epileptic spasms and status epilepticus were prevalent in the activation group (Fig. 1D). Consistently, the activation group also showed a higher prevalence of DEE (Fig. 1E) and severe ID (Fig. 1F). Statistical analysis further confirmed a better response to SCBs in the inactivation group compared with the activation group (χ^2^ tests and Fisher’s exact tests, *P* < 0.0001, Fig. 1G and H), whereas non-SCBs treatments had comparable effects in both groups. Given that most individuals in the activation group had severe DEE, whereas the phenotypic spectrum in the inactivation group was more heterogenous, we further restricted our clinical pharmacological comparison to individuals with DEE and severe ID. Within this population, SCBs treatment remained significantly more effective in the inactivation group (χ^2^ test, *P* = 0.0074, and Fisher’s exact test, *P* = 0.0431, Supplementary Fig. 2A and B). In conclusion, these findings indicate that the activation and inactivation groups constitute two clinically heterogeneous populations that differ in both phenotypic severity and pharmacological response.

### In vitro drug effect on WT Na_V_1.6 channels

To understand why individuals with inactivation-related variants exhibited a better response to SCBs than those with activation-related variants, we conducted a series of pharmacological studies. We selected PHT as the representative drug of classical SCBs, and PRAX-330 as an alternative new generation SCB, since it has been shown to be effective in *SCN8A* DEE variants^18,20^. We first tested the effect of these two drugs on Na_V_1.6 WT channels. To mimic physiological channel composition, we co-expressed Na_V_1.6 α subunit with β1- and β2-subunits in neuroblastoma ND7/23 cells to investigate how PHT and PRAX-330 modulate biophysical properties of Na_V_1.6 channels. Endogeneous Na^+^ channels were then blocked with TTX so that solely the transfected Na_V_1.6 channels were recorded^1,6^ and could be specifically modulated by PHT and PRAX-330.

As PHT binds to the inactivated state of Na_V_ in a relatively slow manner^15,29^, a conditioning pulse lasting 2500 ms at different potentials was applied to allow binding of PHT and PRAX-330, followed by a 25 ms testing pulse to −10 mV. Representative Na^+^ current traces are shown in Fig. 2A. Consistent with previous reports^13,29^, administration of 100µM PHT or 0.5 µM PRAX-330 caused a reduction of Na^+^ currents at −70 mV conditioning pulse but not at more hyperpolarized potentials (e.g.-130 mV, Fig. 2A). Steady-state inactivation curves were obtained by fitting a Boltzmann function to the normalized peak Na^+^ currents plotted versus conditioning potentials in presence and absence of the drugs (Fig. 2B and C). Both drugs caused a hyperpolarizing shift of the steady-state inactivation curve in a dose-dependent manner (Fig. 2B-D). The IC_50_ of PHT was 26.9 µM and the IC_50_ of PRAX-330 was 0.2 µM, indicating a stronger potency by two orders of magnitude of PRAX-330 compared with PHT. The maximal hyperpolarizing shift of the steady-state inactivation curve caused by PRAX-330 was 14.6 mV, which is also larger than the maximal shift caused by PHT (8.8 mV), also suggesting a higher efficacy of PRAX-330.

**Fig. 2:**
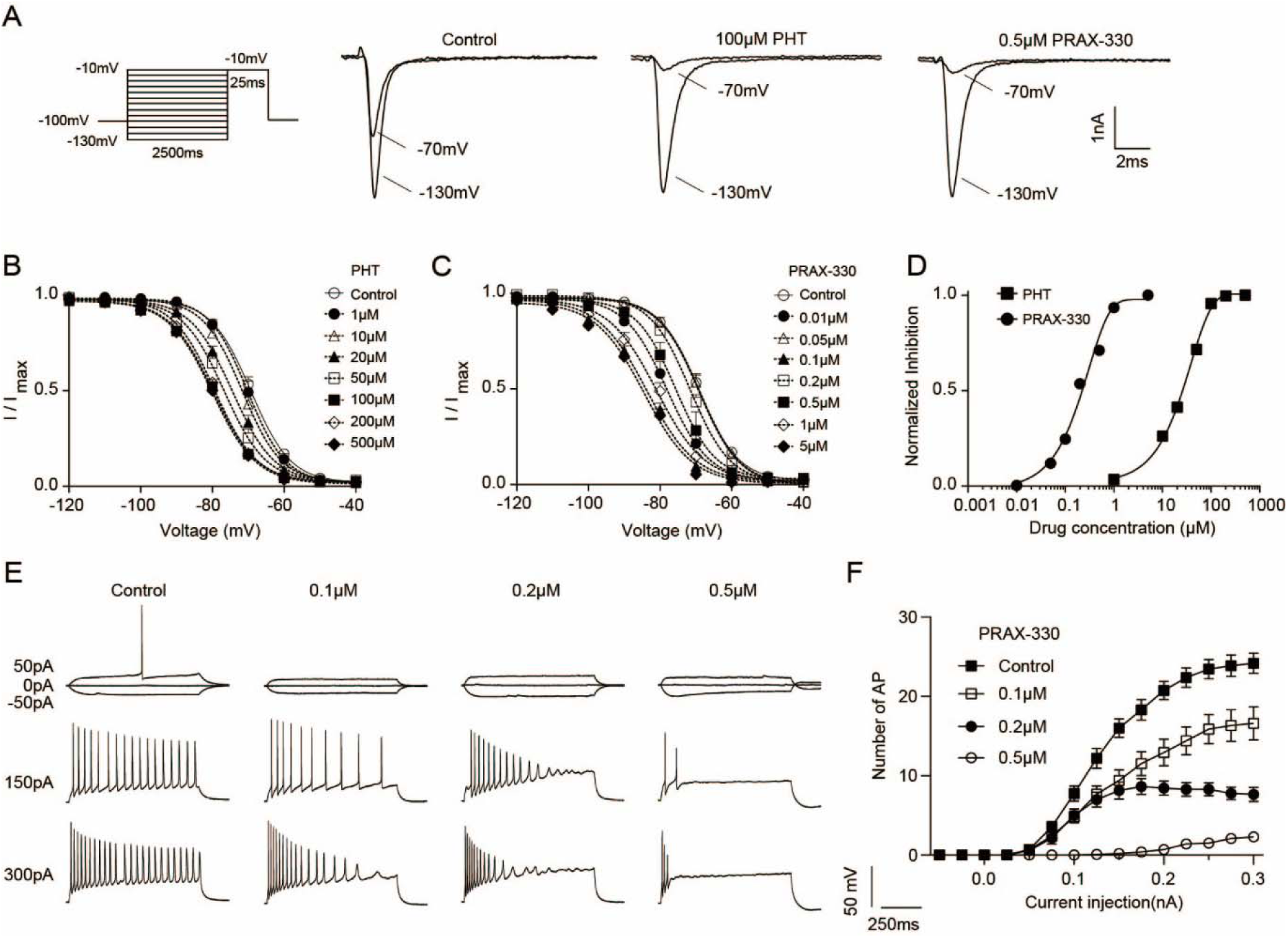
Dose-response relationship of PHT and PRAX-330 in Na_V_1.6 WT channels. (A) Voltage protocol used to examine steady-state inactivation (left) and representative traces of Na^+^ currents evoked by the test pulse to −10 mV following a 2500ms conditioning pulse at −130mV or −70mV recorded in control solution, in the presence of 100 μM PHT or 0.5 μM PRAX-330 respectively. (B) Voltage-dependent steady-state inactivation curves in Na_V_1.6 WT channels at different concentrations of PHT. Lines represent Boltzmann functions fit to the data points. (C) Voltage-dependent steady-state inactivation curves of Nav1.6 WT channels at different concentrations of PRAX-330. Lines represent Boltzmann functions fit to the data points. (D) Dose-response curves for both drugs by plotting normalized shifts of V_1/2_ of the inactivation curve against the concentrations of PHT or PRAX-330. A Hill equation was fit to the data points. (E) In the absence of TTX, representative action potential recordings from neurons expressing Na_V_1.6 WT channels under control conditions or in the presence of different concentrations of PRAX-330. (F) Input–output curves showing the number of action potentials evoked by incremental current injections at different concentrations of PRAX-330.

We next tested the effect of both drugs on neuronal firing in cultured primary mouse hippocampal neurons transfected with WT Na_V_1.6 channels in the absence of TTX. 0.5 μM PRAX-330 almost abolished neuronal firing (Fig. 2E and F), thereby limiting its use in evaluating variant-specific effects on neuronal excitability at this concentration. This may be due to PRAX-330’s additional effect on the inhibition of persistent current and enhancement of use-dependent block. Based on the results obtained from ND7/23 cells and primary neuronal cultures, we set the working concentrations of PHT and PRAX-330 at 100 μM and 0.2 μM respectively, for all subsequent experiments. (Fig. 2D).

### Effect of PHT on biophysical properties of WT and mutant Na_V_1.6 channels

We selected two activation-related (F846S and M1760I) and two inactivation-related (G1475R and N1877S) variants to examine the effects of PHT and PRAX-330 on: (1) tonic current, (2) voltage-dependent steady-state activation and inactivation, (3) persistent current and (4) use-dependent block. We also determined the binding and unbinding kinetics for these two drugs. As for the WT, all mutant α-subunits were transfected together with β1- and β2-subunits into ND7/23 cells and studied under TTX to block all endogeneous Na^+^ channels.

In line with our previous studies^1,6^, all four variants did not affect the peak Na^+^ current amplitude/density compared with WT channels (Fig. 3A, Supplementary Table 2); M1760I and F846S caused a hyperpolarizing shift of the steady-state activation curve (V_1/2 WT_ = −20.8 ± 0.8 mV; V_1/2 M1760I_ = −25.9 ± 1.4 mV, *P* < 0.0001; V_1/2 F846S_ = −30.0 ± 1.1 mV, *P* < 0.0001, one-way ANOVA with Dunnett’s test), whereas G1475R and N1877S caused a depolarizing shift of the steady-state inactivation curve (V_1/2 WT_ = −69.2 ± 0.9 mV; V_1/2 G1475R_ = −62.1 ± 1.1 mV, *P* < 0.0001; V_1/2 N1877S_ = −66.5 ± 1.5 mV, *P* < 0.0001, one-way ANOVA with Dunnett’s test, Fig. 3B and Supplementary Table 2). Applying 100 μM PHT had no relevant effects on peak Na^+^ current amplitude/density or steady-state activation for WT and four variants (Fig. 3A and B). As expected, PHT induced a depolarizing shift of the steady-state inactivation curve, with comparable amplitude for all clones (ΔV_1/2 WT_ = 10.4 mV, ΔV_1/2 M1760I_ = 10.6 mV, ΔV_1/2 F846S_ = 8.9 mV, ΔV_1/2 G1475R_ = 9.9 mV, ΔV_1/2 N1877S_ = 10.5 mV, Fig. 3B and Supplementary Table 2).

**Fig. 3:**
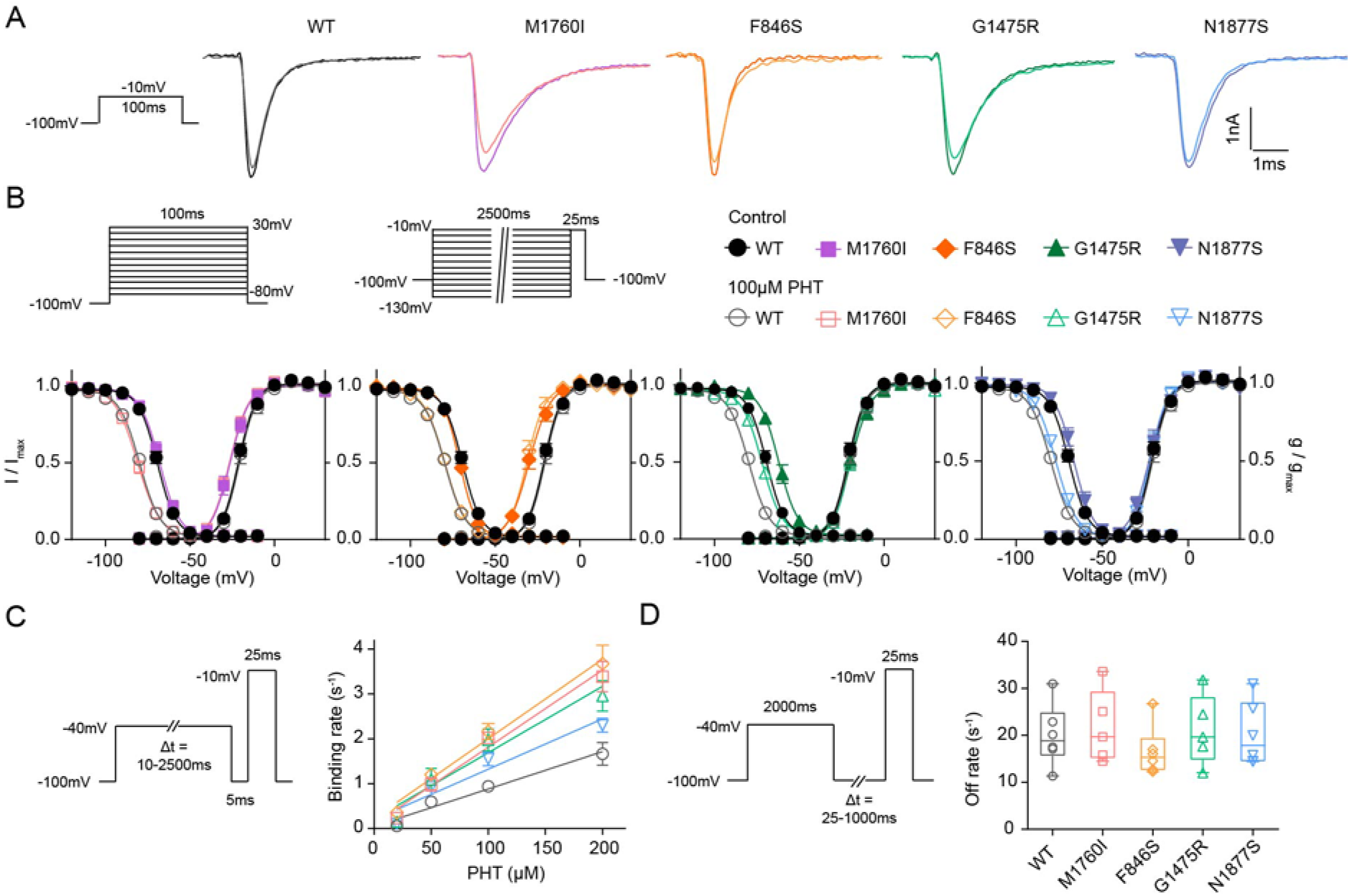
Effect of phenytoin on biophysical properties of WT and mutant Na_V_1.6 channels. (A) Representative Na^+^ current traces elicited by a step depolarization of −10 mV for WT and mutant Na_V_1.6 channels in the absence (dark color) and presence (light color) of PHT. (B) Upper: Voltage protocols for assessing steady-state activation and inactivation kinetics. For steady-state activation (left), Na^+^ currents were elicited by a series of depolarizing sweeps lasting 100 ms ranging from −80 to +30 mV in 10 mV increments, with a 2s interval holding at −100 mV between sweeps. For steady-state inactivation (middle), a 2500 ms conditioning pulse ranging from −130 mV to −10 mV in 10mV increments was applied, followed by a 25 ms test pulse to −10 mV. The symbols are shown on the right. Lower: Voltage-dependent steady-state activation and inactivation curves of WT and four variants, respectively. Lines represent Boltzmann functions fit to the data points. Data are presented as means ± SEM. (C) Left: Voltage protocol for examining binding kinetics of PHT. Right: The mean binding rates of PHT for WT and four variants plotted against PHT concentration, which were fitted by a linear regression to decide the slope as the K_on_. Pre-pulse durations were 10, 20, 50, 100, 500, 1000, 1500 and 2500 ms respectively. Data are presented as means ± SEM. (D) Left: Voltage protocol for assessing unbinding kinetics of PHT. Right: Unbinding rates of PHT for WT and four variants. Recovery durations were 25, 50, 100, 250, 500 and 1000ms respectively. Data are shown as box-and-whisker plot with the 25^th^; 50^th^ and 75^th^ percentiles, minima and maxima. Individual data points are also displayed. Detailed statistical analysis is provided in Supplementary Table 2.

Notably, M1760I exhibited a pronounced increased persistent current, accounting for around 4% of the peak Na^+^ current at +10 mV (Supplementary Fig. 3A). PHT failed to suppress this increased persistent current in M1760I (upon a step depolarization of +10mV lasting 100 ms, I_persistent control_ = 3.8 ± 0.2% vs. I_persistent PHT_ = 3.3 ± 0.3%, *P* = 0.2247, two-way ANOVA with post hoc Sidak’s test, Supplementary Table 2). Persistent currents in WT and other variants remained comparable in the absence and presence of 100 µM PHT (Supplementary Table 2). Among the four variants, only G1475R showed less use-dependent current reduction at 80Hz compared with WT in the absence of PHT. Applying 100 µM PHT led to a strong use-dependent block for G1475R, WT and all other mutants, reducing the Na^+^ current to similar amounts (Supplementary Fig. 3B).

As the dynamic interaction between Na_V_ channels and their blockers can significantly influence the effect of drug action, we determined the binding and unbinding rates of PHT using the voltage-step protocols illustrated in Fig. 3C and D. Representative Na^+^ current traces and the determination of binding and unbinding rates for WT channels were shown in Supplementary Fig. 1 (for details see Supplementary materials and methods). The binding and unbinding rates of PHT for mutant channels were assessed using the same approach. The binding rate constants were determined as the slope of a linear regression fit to the binding rates plotted against PHT concentrations (Supplementary Fig. 1D). Interestingly, the two activation-related variants exhibited the highest PHT binding rate constants, the two inactivation-related variants showed intermediate binding rate constants, and WT channels showed the lowest binding rate constant (K_on WT_ = 8.3 ± 1.2 x 10^3^ M^-1^ s^-1^, K_on M1760I_ = 17.2 ± 1.4 x 10^3^ M^-1^ s^-1^, K_on F846S_ = 17.7 ± 1.8 x 10^3^ M^-1^ s^-1^, K_on G1475R_ = 14.8 ± 2.1 x 10^3^ M^-1^ s^-1^, K_on N1877S_ = 11.2 ± 1.3 x 10^3^ M^-1^ s^-1^, Fig.3C and Supplementary Table 2). However, statistical significance was observed only for the two activation-related variants compared with WT (K_on WT_ vs. K_on M1760I_, *P* = 0.0077; K_on WT_ vs. K_on F846S_, *p* = 0.0043, one-way ANOVA with Dunnett’s test, Supplementary Table 2). In contrast, the unbinding kinetics of PHT showed no significant differences between WT and the four variants (K_off WT_ = 20.0 ± 2.7 s^-1^, K_off M1760I_ = 21.7 ± 3.5 s^-1^, K_off F846S_ = 16.6 ± 2.2 s^-1^, K_off G1475R_ = 21.1 ± 3.3 s^-1^, K_off N1877S_ = 20.2 ± 2.8 s^-1^, Fig. 3D and Supplementary Table 2). This suggests that while activation-related variants may modulate channel conformation to favor PHT binding, they may not substantially alter the stability of the drug-channel complex once it is formed.

### Effect of PHT on action potential firing in neurons expressing WT or mutant Nav1.6 channels

Our results in ND7/23 cells showed that PHT inhibited the activity of WT and mutant Na_V_1.6 channels mainly by causing a hyperpolarizing shift of steady-state inactivation and enhancing the use-dependent block. PHT thus corrected the main defect of inactivation-related variants, whereas the main defect of the activation-related variants, i.e. the hyperpolarizing shift of the activation curve, remained unaltered by the drug, which may explain the differential response of both variant groups to classical SCBs. To corroborate this hypothesis, we therefore transfected WT or mutant Na_V_1.6 channels into primary cultured mouse hippocampal neurons and examined action potential firing properties in transfected neurons in the absence and presence of the drugs to further evaluate their neurophysiological impact. In transfected neurons, action potential firing was jointly driven by the transfected WT or mutant Na_V_1.6 and endogenous Na^+^ channels in the absence of TTX, partially simulating the heterozygous situation of variants in affected individuals, as has been shown as a valid model previously by our group^1,6,17^.

A series of current injections ranging from −50 to 300 pA in 25 pA increments elicited action potential firing and representative traces are shown in Fig. 4A. The input-output relationships (Fig. 4B) and analysis of the area under the input-output curves (AUC) (Fig. 4C) indicated that all four GOF variants did not have a significant effect on action potential firing compared with WT in the absence of PHT. However, M1760I and F846S exhibited a significant lower firing threshold (WT: −41.8 ± 0.8 mV; M1760I: −45.5 ± 0.9 mV, *P* = 0.006; F876S: −46.8 ± 0.8 mV, *P* < 0.0001, Two-way ANOVA with post hoc Sidak’s test, Fig. 4D and Supplementary Table 3), consistent with our previous findings^1,6^.

**Fig. 4:**
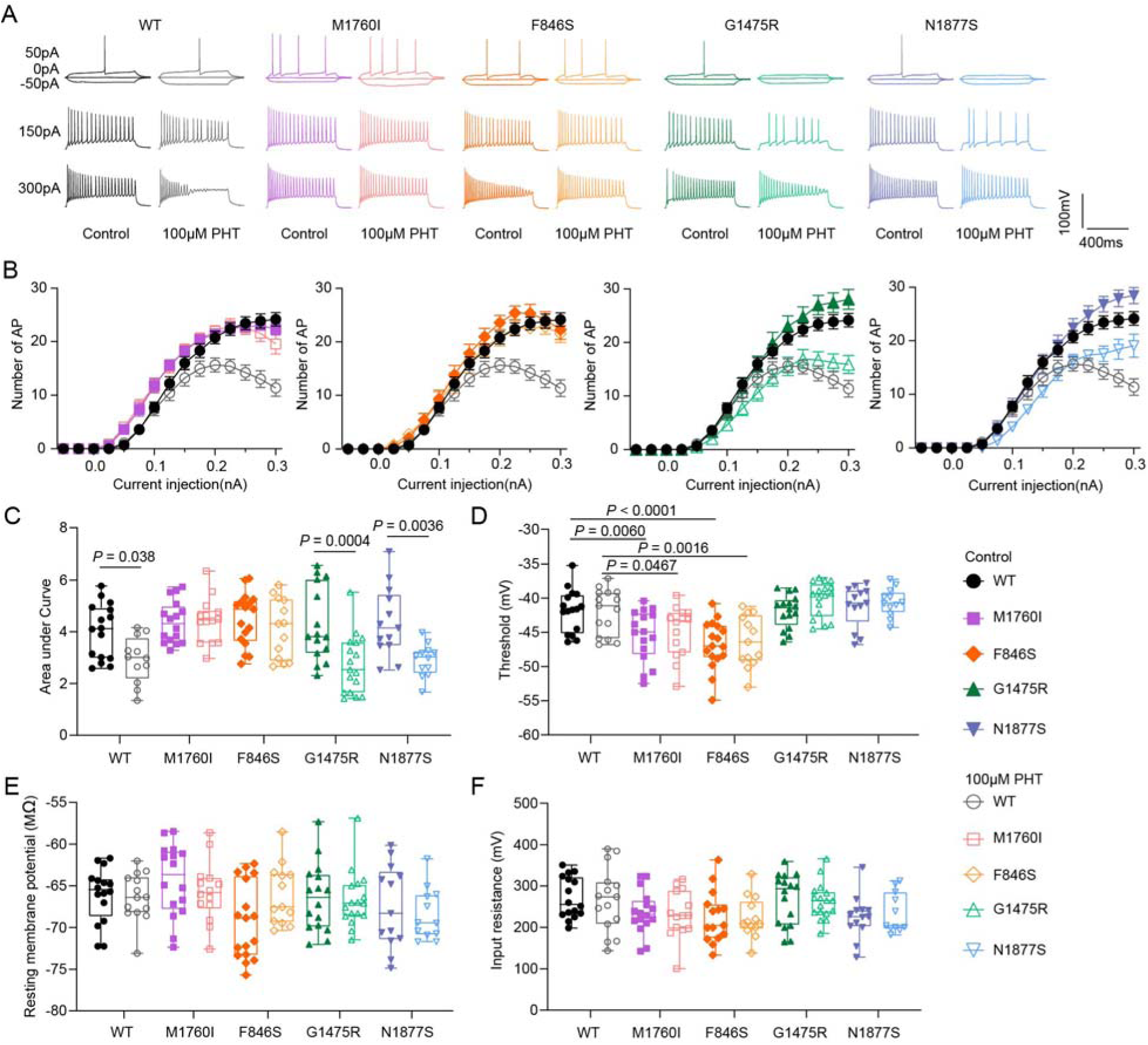
Effect of phenytoin on neuronal firing in neurons expressing WT or mutant Na_V_1.6 channels. (A) Representative traces of action potentials recorded in the neurons transfected with WT or mutant Na_V_1.6 channel variants in the absence (dark color) and presence of (light color) 100 µM PHT. (B) Number of action potentials (AP) plotted versus injected current for WT and four variants. Data are presented as means ± SEM. (C) Area under the curve (AUC) for the input-output relationships shown in (B). (D) Threshold of the first evoked action potential. (E) Resting membrane potential and (F) Input resistance of recorded neurons. Two-way ANOVA with Sidak post hoc test were performed. Data in panel (C) to (F) are shown as box-and-whisker plot with the 25^th^; 50^th^ and 75^th^ percentiles, minima and maxima. Individual data points are also displayed. Detailed statistical analysis is provided in Supplementary Table 3.

Application of 100 µM PHT markedly reduced action potential firing in neurons expressing WT, G1475R or N1877S revealed by a significant decrease of the AUC (two-way ANOVA with post hoc Sidak’s test, Fig. 4C, Supplementary Table 3). Interestingly, PHT had no effect on the action potential firing in neurons expressing M1760I or F846S as the AUC remained comparable in the absence and presence of 100 µM PHT (Fig. 4C, Supplementary Table 3). The increased threshold in F846S or M1760I expressing neurons were also not restored by PHT (Fig. 4D, Supplementary Table 3). Passive neuronal properties such as resting membrane potential and input resistance were unaffected by either the variants or PHT (Fig. 4E and F, Supplementary Table 3). These results fit well with and can explain the clinical data that individuals with inactivation-related variants responded better to SCBs (e.g. PHT) than those with activation-related variants. Considering that PHT affected inactivation but not activation gating of Na_V_1.6 channels in ND7/23 cells, it is conceivable that the biophysical defect of the activation-related variants cannot be addressed by classical SCBs, and despite an effective use-dependent block (Supplementary Fig. 3) PHT is not effective in reducing neuronal firing for these variants. This correlates nicely with its limited therapeutic efficacy in treating individuals with activation-related *SCN8A* GOF variants.

### Effect of PRAX-330 on biophysical properties of WT and mutant Na_V_1.6 channels

PRAX-330 is a new SCB with much higher affinity to Na^+^ channels which has been shown to effectively attenuate abnormal neuronal activity, seizures and reduced survival in two *Scn8a* DEE mouse models^18–20^. Similar to 100 µM PHT, the peak Na^+^ current amplitude of either WT or mutant Na_V_1.6 channels is unaffected in the presence of 0.2 µM PRAX-330 (Fig. 5A, Supplementary Table 4). Using the same voltage protocol as for assessing the effects of PHT on activation kinetics (2s interval between sweeps holding at −100 mV), we found that PRAX-330 induced a small depolarizing shift of the steady-state activation curve as well as a reduced slope (shown as light dashed lines in Fig. 5B, Supplementary Table 4). The amplitude of this shift was comparable across WT and all four variants (ΔV_1/2 WT_ = 3.6 mV, ΔV_1/2 M1760I_ = 2.3 mV, ΔV_1/2 F846S_ = 3 mV, ΔV_1/2 G1475R_ = 3.1 mV, ΔV_1/2 N1877S_ = 4.3 mV, Fig. 5A and Supplementary Table 4). However, when the interval between sweeps of the voltage protocol was extended from 2s to 10s, the depolarizing shift and reduced slope in the presence of PRAX-330 were no longer observed (shown as light solid lines in Fig. 5B). Furthermore, the presence of 0.2 µM PRAX-330 caused a hyperpolarizing shift of the steady-state inactivation curve (Fig. 5B, Supplementary Table 4), with similar amplitudes in WT and all four mutant channels (ΔV_1/2 WT_ = 8.8 mV, ΔV_1/2 M1760I_ = 7.7 mV, ΔV_1/2 F846S_ = 8.9 mV, ΔV_1/2 G1475R_ = 7.5 mV, ΔV_1/2 N1877S_ = 7.5 mV, Fig. 5B and Supplementary Table 4).

**Fig. 5:**
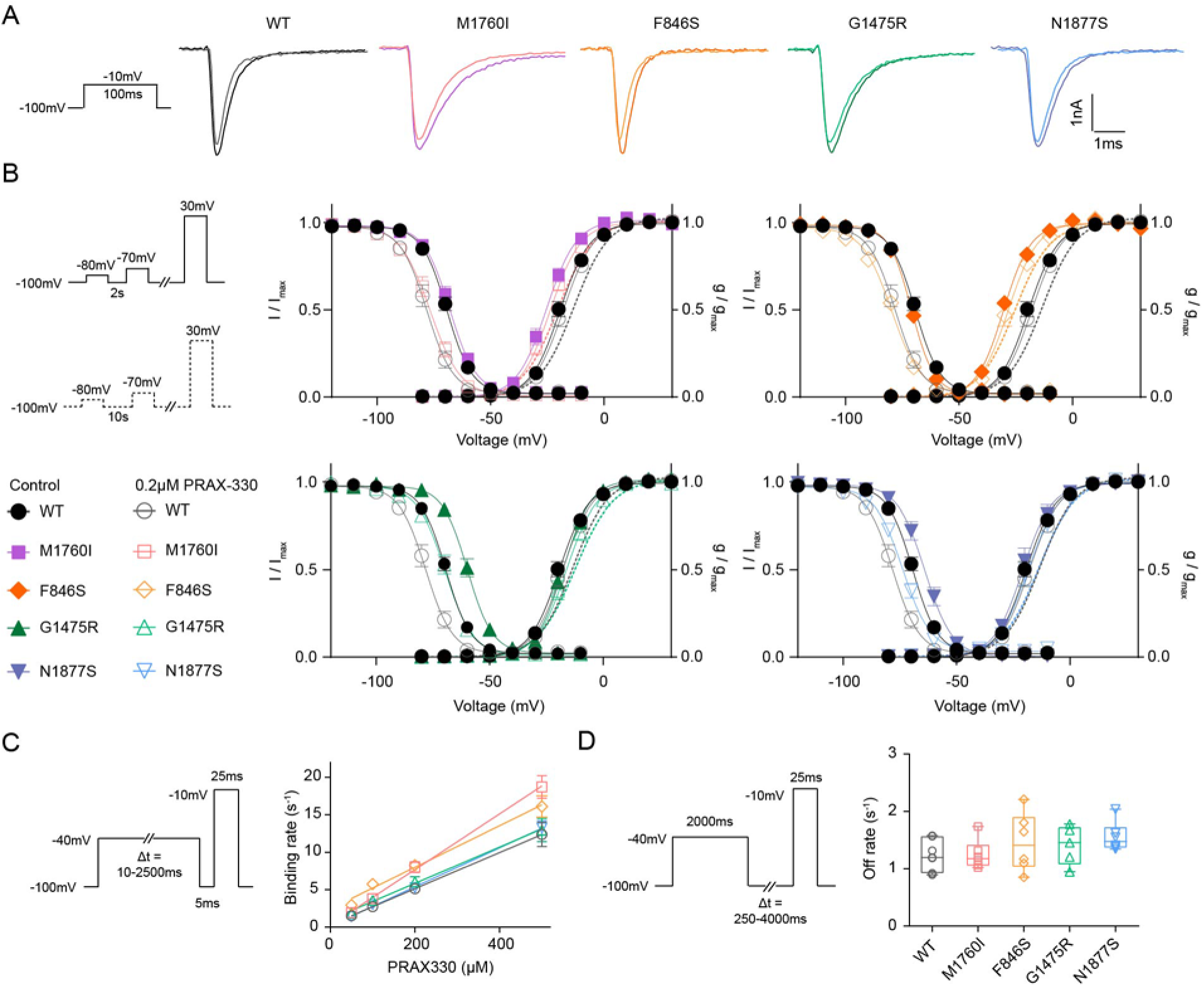
Effect of PRAX-330 on biophysical properties of WT and mutant Na_V_1.6 channels. (A) Representative Na^+^ current traces elicited by a step depolarization of −10 mV for WT and mutant Na_V_1.6 channels in the absence (dark color) and presence (light color) of PRAX-330. (B) Left: Voltage protocols for assessing steady-state activation kinetics. Na^+^ currents were elicited by a series of depolarizing sweeps lasting 100 ms ranging from −80 mV to +30 mV in 10 mV increments, with a 2s (solid line) or 10s (dashed line) interval holding at −100 mV between sweeps. Except for the different hyperpolarization intervals, general protocols for steady-state activation and inactivation kinetics are same as in Figure 3B. For steady-state activation (left), The symbols are shown on the lower side. Right: Voltage-dependent steady-state activation (2 s interval in solid line and 10 s interval in dashed line) and inactivation curves of WT and four variants, respectively. Lines represent Boltzmann functions fit to the data points. Data are presented as means ± SEM. (C) Left: Voltage protocol for examining binding kinetics of PRAX-330. Right: The mean binding rates of PRAX-330 for WT and four variants plotted against PRAX-330 concentration, which were fitted by a linear regression to decide the slope as the K_on_. Pre-pulse durations were 10, 20, 50, 100, 500, 1000, 1500 and 2500 ms respectively. Data are presented as means ± SEM. (D) Left: Voltage protocol for assessing unbinding kinetics of PRAX-330. Right: Unbinding rates of PRAX-330 for WT and four variants. Recovery durations were 25, 50, 100, 250, 500 and 1000ms respectively. Data are shown as box-and-whisker plot with the 25^th^; 50^th^ and 75^th^ percentiles, minima and maxima. Individual data points are also displayed. Detailed statistical analysis is provided in Supplementary Table 4.

In contrast to PHT, PRAX-330 significantly inhibited the increased persistent current in M1760I (Upon a depolarization of +10 mV lasting 100 ms, I_Persistent Control_: = 3.8 ± 0.2% vs. I_Persistent PRAX-330_: 2.2 ± 0.3%, *P* = 0.0002, Two-way ANOVA with post hoc Sidak’s test; Supplementary Fig. 4A). However, similar to PHT, PRAX-330 also did not affect the persistent currents in WT and the other three variants (Supplementary Fig. 4A and Supplementary Table 4). Additionally, upon a 20 Hz depolarization stimulation, PRAX-330 enhanced the use-dependent block in both WT and four variants.

Binding and unbinding kinetics of PRAX-330 were examined using the same protocols as for PHT (Fig. 5C and D). Strikingly, compared with PHT, PRAX-330 showed remarkably faster binding kinetics in Na_V_1.6 WT channels revealed by the approximately 3000 times higher binding rate constant (PRAX-330: K_on WT_ = 24.1 ± 1.9 x 10^6^ M^-1^ s^-1^; PHT: K_on WT_ = 8.3 ± 1.2 x 10^3^ M^-1^ s^-1^). The binding of PRAX-330 to the four mutant channels was also much faster than that of PHT (Supplementary Table 2 and 4). Particularly, M1760I exhibited the highest binding rate constant (K_on M1760I_ = 37.2 ± 2.5 x 10^6^ M^-1^ s^-1^, K_on F846S_ = 27.8 ± 2.5 x 10^6^ M^-1^ s^-1^, K_on G1475R_ = 24.2 ± 2.4 x 10^6^ M^-1^ s^-1^, K_on N1877S_ = 26.1 ± 1.5 x 10^6^ M^-1^ s^-1^, Fig. 5C, Supplementary Table 4). In addition to its fast binding rate, PRAX-330 dissociated relatively slowly from Na_V_ channels, with an off-rate ranging from 1.2-1.6 s^-1^ in WT and mutant channels (K_off WT_ = 1.24 ± 0.10 s^-1^, K_off M1760I_ = 1.25 ± 0.11 s^-1^, K_off F846S_ = 1.46 ± 0.21 s^-1^, K_off G1475R_ = 1.41 ± 0.15 s^-1^, K_off N1877S_ = 1.56 ± 0.11 s^-1^, Fig. 5D, Supplementary Table 4). These distinct pharmacokinetic properties of PRAX-330 may contribute to its higher potency and efficacy compared with PHT (Fig. 2).

### Effect of PRAX-330 on action potential firing in neurons expressing WT or mutant Nav1.6 channels

We next examined the effect of 0.2 µM PRAX-330 on neuronal firing using the same approach as for PHT. Overall, PRAX-330 exhibited a stronger inhibitory effect on neuronal firing compared with PHT, characterized by the presence of a rapid and robust depolarizing block at high current injections for both WT and mutant channels, which are absent in neurons treated with PHT (Fig. 4A and 6A). Analysis of the AUC confirmed a significant reduction in action potential firing in neurons expressing WT and mutant Na_V_1.6 channels to comparable levels (two-way ANOVA with post hoc Sidak’s test, Fig. 6B and C, Supplementary Table 5). Interestingly, PRAX-330 induced a much larger reduction of the AUC than PHT for WT and all four variants (Fig. 4C and 6C, Supplementary Table 3 and 5). Additionally, in neurons expressing M1760I mutant channels, PRAX-330 increased the action potential threshold (M1760I_Control_: −45.5 ± 0.9 mV vs. M1760I_PRAX-330_: −40.1 ± 0.6 mV, *P* < 0.0001, two-way ANOVA with post hoc Sidak’s test, Fig.6D, Supplementary Table 5). The threshold for WT and the other three variants also showed a tendency towards an increase in the presence of PRAX-330, not reaching statistical significance (Supplementary Table 5). PRAX-330 did not alter the resting membrane potential or input resistance in neurons expressing WT or mutant channels (Fig. 6E and F, Supplementary Table 5).

**Fig. 6:**
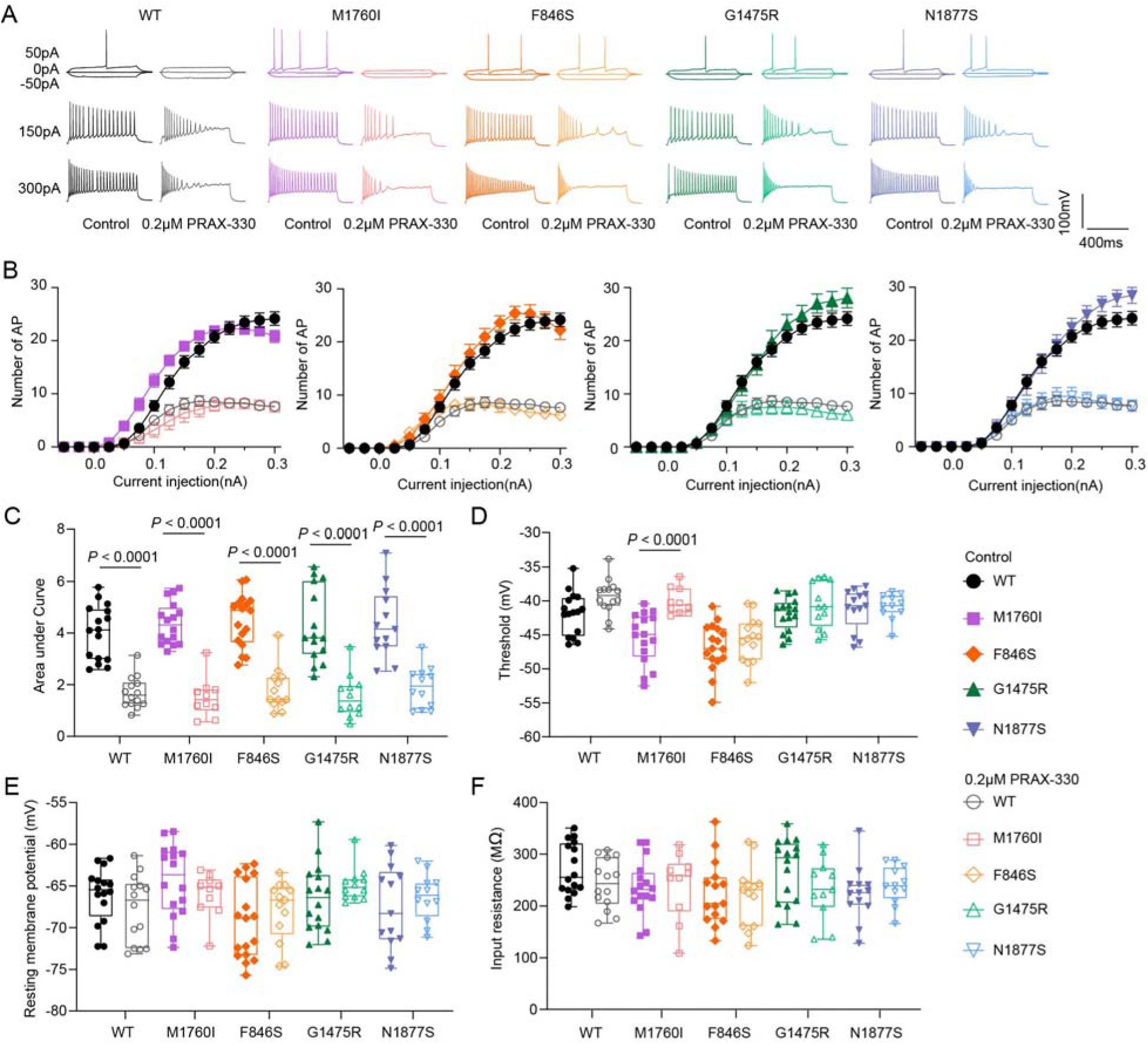
Effect of PRAX-330 on neuronal firing in neurons expressing WT or mutant Na_V_1.6 channels. (A) Representative action potentials recordings in primary cultured mouse neurons transfected with WT or mutant Na_V_1.6 channel in the absence (dark color) or presence of (light color) 0.2 µM PRAX-330. (B) Number of action potentials (AP) plotted versus injected current for WT and all four variants. Data are presented as means ± SEM. (C) Area under the curve (AUC) for the input-output relationships shown in (B). (D) Threshold of the first evoked action potential. (E) Resting membrane potential and (F) Input resistance of recorded neurons. Two-way ANOVA with Sidak post hoc test was performed. Data in panel (C) to (F) are shown as box-and-whisker plot with the 25^th^; 50^th^ and 75^th^ percentiles, minima and maxima. Individual data points for each recorded cell are also displayed. Detailed statistical analysis is provided in Supplementary Table 5.

The strong inhibitory effect of PRAX-330 on action potential firing in neurons expressing activation-related variants fits well with previous reports that PRAX-330 is effective to restore abnormal neuronal activities in *Scn8a* DEE mice^18–20^. The much stronger effects of PRAX-330 in comparison to PHT suggest that this new SCB – or similar derivatives – may have the potential to overcome resistance of GOF *SCN8A* variant carriers to classical SCBs, including those causing primarily activation defects.

## Discussion

In this study, we compared the clinical phenotypes and pharmacological responses of 173 affected individuals with two major classes of *SCN8A* GOF variants, namely those that alter primarily either activation or inactivation gating of Na_V_1.6 channels. We found that activation-related variants generally cause more severe epileptic phenotypes with earlier seizure onset, which are often refractory to classical SCBs with approximately one-third of affected individuals showing no clinical benefit. In contrast, *SCN8A* inactivation-related variants lead to milder forms of epilepsy with a better response to SCBs. To understand the differential clinical pharmacological response, we conducted experimental pharmacological studies in both neuroblastoma cells and neuronal cultures to examine the effect of PHT (representing classical SCBs) and PRAX-330 (representing new SCBs) on two activation-related (F846S and M1760I) and two inactivation-related variants (G1475R and N1877S). Consistent with clinical observations, PHT suppressed action potential firing in neurons expressing the two inactivation-related variants but not those expressing the two activation-related variants. Along our hypothesis, it is conceivable that this effect can be attributed to the mechanism of action of classical SCBs, which induce hyperpolarizing shift of the steady-state inactivation curve, thus correct one of the main biophysical defects of the inactivation-related variants, but do not affect activation gating so that the main defect of the activation-related variants, the hyperpolarizing shift of the activation curve, remains unchanged. The effect of PHT on the inactivation curve and use-dependence, which was also visible in the activation-related variants, was obviously not able to overcome the persisting activation defect, as neuronal firing could not be significantly reduced for the activation related variants.

In contrast, the new SCB PRAX-330 showed greater efficacy than PHT revealed by its ability to inhibit neuronal firing for both activation- and inactivation-related variants. Our results strongly suggest, that this results from the alterations of both activation and inactivation gating induced by PRAX-330, which were enabled by its much faster binding and moderately slower unbinding kinetics to/from both WT and mutant Na_V_1.6 channels compared to PHT. We therefore conclude that (i) severe biophysical alterations caused by *SCN8A* activation-related variants can lead to drug-resistant epilepsy; (ii) PRAX-330 (and related compounds with similar mechanisms of action) may represent new and promising therapeutic options.

Since the first case of *SCN8A*-related epilepsy was reported^24^, >260 (likely) pathogenic missense variants have been registered on ClinVar, more than half of which were associated with drug-resistant developmental and epileptic encephalopathy^30,31^. However, only a limited number of these variants has been functionally characterized^1,32^. Previously, by assessing neuronal firing using a single-compartment conductance-based model, we found that the changes of activation gating caused a strong effect on neuronal firing, while alterations of peak Na^+^ current showed a moderate effect and the changes of inactivation kinetics or persistent current had a mild effect on neuronal firing^1^. Combining the functional results with clinical data, we demonstrated that the clinical severity of epilepsies caused by *SCN8A* GOF variants correlates with a score of biophysical changes induced by the corresponding variants. Variants that had stronger GOF effects on gating parameters were predominantly associated with more severe epilepsy syndromes.^1,6,33^. The current studies (i) support our previous findings by showing that activation-related variants were associated with more severe clinical phenotypes than inactivation-related variants; (ii) expand the previous correlation by indicating that the responses of affected individuals with *SCN8A* GOF variants to SCBs at least also partially correlated with the biophysical changes of variants; (iii) highlight the importance of comprehensive functional and pharmacological investigations for pathogenic *SCN8A* variants in both heterologous expression systems and neuronal cultures to guide clinical treatment: classical SCBs are likely to be effective for the treatment of individuals with inactivation-related variants, whereas individuals with activation-related variants may be refractory to classical SCBs thus require novel treatments with different pharmacological properties, as shown here for PRAX-330.

PRAX-330 was originally developed as an anti-arrhythmic agent and characterized for its potent suppression of late/persistent sodium current in Na_V_1.5 channels^29,34,35^. As similar inhibitory effects on persistent current have been observed on other subtypes of Na_V_ channels (e.g. Na_V_1.1, Na_V_1.2 and Na_V_1.6), PRAX-330 has shown strong anticonvulsant efficacy in multiple mouse models, reducing seizures and preventing premature death in genetically modified mice such as *Scn1a*^L1649Q/+^, *Scn2a*^Q54^, *Scn8a*^N1768D/+^ and *Scn8a*^R1872W/+^ ^18–20,36,37^. The suppression of late/persistent/ramp current has been considered as the principal mechanism of PRAX-330’s anti-seizure action^38^. In our current studies, M1760I increased the persistent current that was only inhibited by PRAX-330 but not PHT, which may contribute to the effect on neuronal firing for this variant, but not for the other three, which did not show an increased persistent current compared to the WT (and no modulation of the small persistent current by PRAX-330). Yet, PRAX-330 still robustly suppressed neuronal firing for these variants. PRAX-330 (0.2 µM) also induced a hyperpolarizing shift of the inactivation curve with similar amplitudes across WT and all four variants, which are even slightly smaller than those induced by PHT (100 µM). However, PHT failed to suppress neuronal firing for activation-related variants. The effect of 0.2 µM PRAX-330 on persistent current and inactivation gating thus probably contributes to but does not fully explain the potent inhibitory effects of PRAX-330 on action potential firing in neurons expressing activation-related variants.

We therefore quantified the binding and unbinding kinetics of both PHT and PRAX-330 on Na_V_1.6 α subunits co-expressed with β1 and β2 subunits in ND7/23 cells. The binding rate constant and unbinding rate of PHT observed in our experiments are consistent with previous reports showing slow binding and fast unbinding kinetics^13,39^. Compared with PHT (and other classical SCBs such as CBZ and LTG^13^), PRAX-330 showed approximately 3000 times faster binding and moderately slower unbinding kinetics from WT and mutant Na_V_1.6 channels, similar to the findings for PRAX-330 in human induced pluripotent stem cell-derived cardiomyocytes^40^. These kinetic differences between PRAX-330 and PHT likely underlie their divergent efficacy. The super-fast binding and moderately slow unbinding of PRAX-330 suggest that this drug binds highly rapidly as soon as the channels open and inactivate, prolonging occupancy of the inactivated state and leading to an indirect accumulative effect on activation gating. This is reflected by our results showing that the depolarizing shift of the activation curve induced by PRAX-330 was only induced when the intervals (holding at −100 mV) between sweeps were 2 s (Fig. 5B and Supplementary Table 4). When the intervals were extended to 10s, which is sufficient for PRAX-330 to dissociate from Na_V_1.6 channels (K_off_ ≈ 1.2 s ¹), the depolarizing shift of activation curve was not observed anymore. Since PHT binds slowly (hundreds of milliseconds) and dissociate quickly (K_off_ ≈ 20 s ¹) from the inactivated channels, it did not cause the accumulative effect on the activation kinetics (Fig. 3B and Supplementary Table 2), which might limit its efficacy for treating severe epilepsies. We thus expect, that PRAX-330 will be highly effective also for activation-related variants, since it blocks channels much more effectively at high-frequency firing, including an indirect correction of the hyperpolarizing shift of the activation curve.

While PRAX-330 has shown efficacy in pre-clinical models, it is currently still in phase I clinical trials (ACTRN12617001512314), with no efficacy data reported to date. The structurally and mechanistically related compounds Relutrigine (PRAX-562)^41^ and Vormatrigine (PRAX-628)^39^ have both demonstrated robust efficacy in clinical phase II trials in patients with *SCN2A*- and *SCN8A*-related DEE. While their clinical outcomes differ in magnitude, both compounds exhibit rapid binding kinetics, moderate unbinding rates, and preferential suppression of persistent Na^+^ current. These clinical successes collectively support the hypothesis that a balance between binding and unbinding kinetics, together with inhibition of pathological persistent current, may represent a critical determinant of effective new generation, small-molecule SCBs. Recently, a specific blocker of Na_V_1.6 channels, Zandatrigine (NBI-921352)^42^ failed to demonstrate meaningful reduction of seizures and to meet its phase II trials endpoint. Characterizations of the pharmacokinetics for this compound may therefore provide important insights into the rational design of future Na_V_1.6-selective inhibitors.

In summary, our studies further categorized the affected individuals with *SCN8A* GOF variants, showing that those with inactivation–related variants have mild epileptic syndromes and respond better to classical SCBs (e.g. PHT) than those with activation-related variants. PRAX-330 but not PHT inhibited neuronal firing in neurons expressing activation-related variants, enabled by much faster binding and moderate slower unbinding kinetics of PRAX-330 compared with PHT. Novel SCBs with similar binding and unbinding kinetics as PRAX-330 may offer new ‘precision’ therapeutic options for drug-resistant epilepsy caused by *SCN8A* activation-related variants. Our studies highlight the importance of functional characterizations of *SCN8A*-DEE associated variants, provide new insights into drug-resistant epilepsy and lay a theoretical foundation for understanding the mechanisms of action of next-generation SCBs.

## Data availability

All detailed clinical data are available in Supplementary Table 1. Functional and pharmacological data will be made available from the lead contact upon any reasonable request from qualified investigators.

## Acknowledgement

Conception and design of the study: YL, HoL and HaL. Acquisition and analysis of data: HaL, SL, RP, KJ, BG, EG, RM. Writing-original draft: HaL, YL. Writing-review and editing: HaL, YL, HoL, CB.

## Funding

This work was supported by the Federal Ministry for Research, Technology and Space (BMFTR, Treat-ION, 01GM2201A to YL and HoL), by the German Research Foundation (DFG, Research Unit FOR-2715, Le1030/15-2 to HoL), by the foundation ‘no epilep’ (to HoL), by the Else-Kröner-Fresenius Foundation (EKFS, clinician scientist college Precise.net to HoL and CB), and by the DFG and the Medical Faculty Tübingen (MINT Clinician Scientist program to CB, DFG, 493665037).

## Competing Interest

R.S.M. received honoraria from Angelini, Eisai, Immedica, Jazz Pharma, Orion, Saniona and UCB. H.L. received research support from Bial, Boehringer Ingelheim, and Lario Therapeutics; he is a consultant for Lario and Praxis Precision Medicine, and a former consultant for Angelini, Bial, Eisai, and UCB. The remaining authors report no competing interests.

---

**Supplementary Figure 1:**
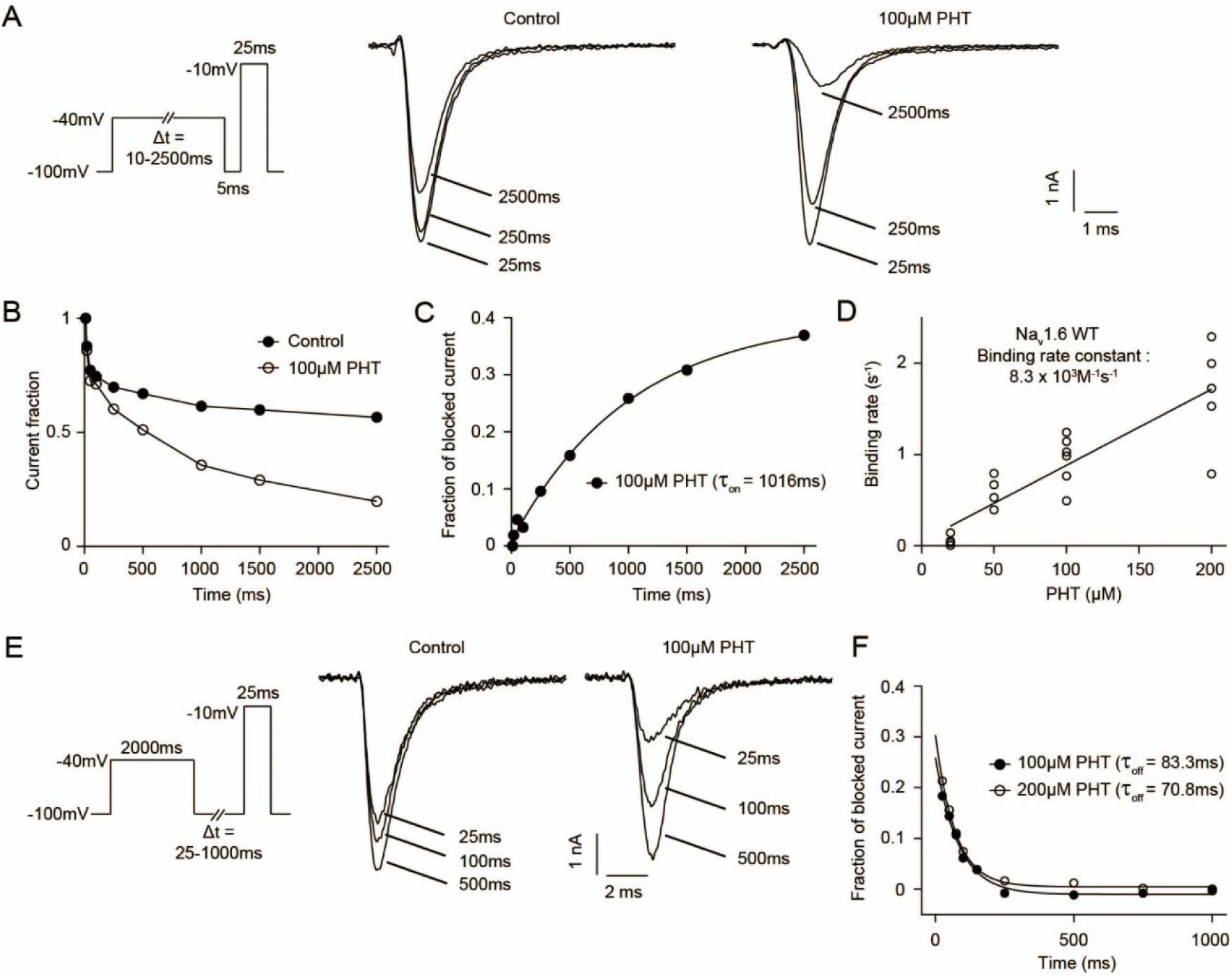
The binding and unbinding kinetics of phenytoin for Na_V_1.6 WT channel. (A) Voltage-clamp protocol and representative traces of Na^+^ currents evoked by the test pulse to −10 mV following a 25, 250 or 2500 ms conditioning pulse to −40 mV and a short hyperpolarizing pulse for recovery from fast inactivation, both in the absence (control) and the presence of 100 μM PHT. (B) Normalized Na^+^ currents from a single cell in the absence and presence of 100 μM PHT plotted versus the pre-pulse duration time. (C) The relatively blocked Na^+^ current was derived from (B), which was determined by subtracting the normalized current recorded in the presence of PHT from the normalized current recorded in control solution. A first-order exponential function was fit to the data points, which were plotted against the pre-pulse duration, to determine the time constant (τ_on_) for development of block in the presence of 100 μM PHT. (D) Binding rates (1/τ_on_, s^−1^) in individual cells expressing WT Na_V_1.6 channels are plotted against the PHT concentrations. The slope of the linear regression provides an estimate of the binding rate constant. (E) Voltage-clamp protocol and representative traces of Na^+^ currents evoked by a 25 ms test pulse to −10 mV, following a 2000 ms conditioning pulse to −40 mV to allow full drug binding and 25, 100 or 500 ms hyperpolarizing pulses to −100 mV for recovery, both in the absence and presence of 100 μM PHT. (F) The relatively blocked current from a single cell was determined by subtracting the normalized current recorded in the presence of PHT from the normalized current recorded in control solution. A first-order exponential function was fit to the data points, which were plotted against the duration of the recovery pulses, to determine the time constant (τ_off_) of PHT unbinding. The similar time constants at 100 or 200 µM PHT showing that the unbinding rate is independent of the PHT concentration.

**Supplementary Fig. 2:**
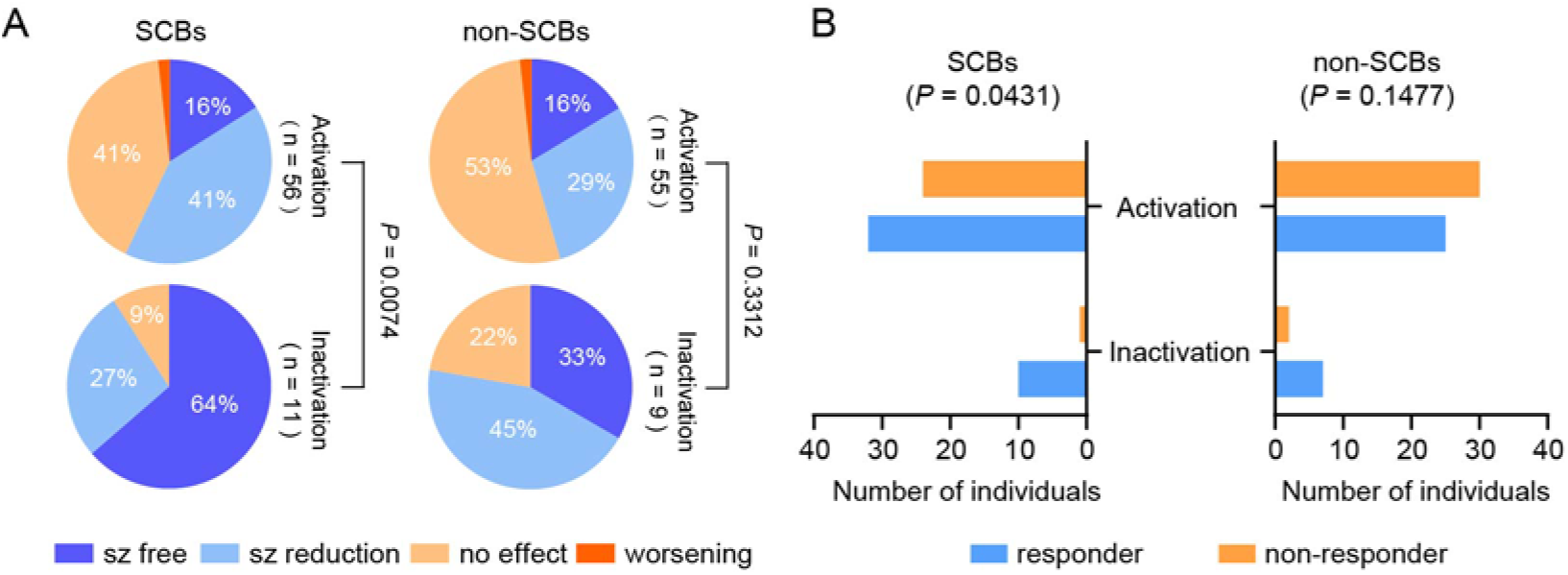
Pharmacological responsiveness of individuals with severe DEE and ID. (A) and (B) showed the treatment response to SCBs and non-SCBs in individuals with DEE and severe ID. P values derived from χ2 tests are provided under the pie chart or from Fisheŕs exact tests are provided above the bar chart.

**Supplementary Fig. 3:**
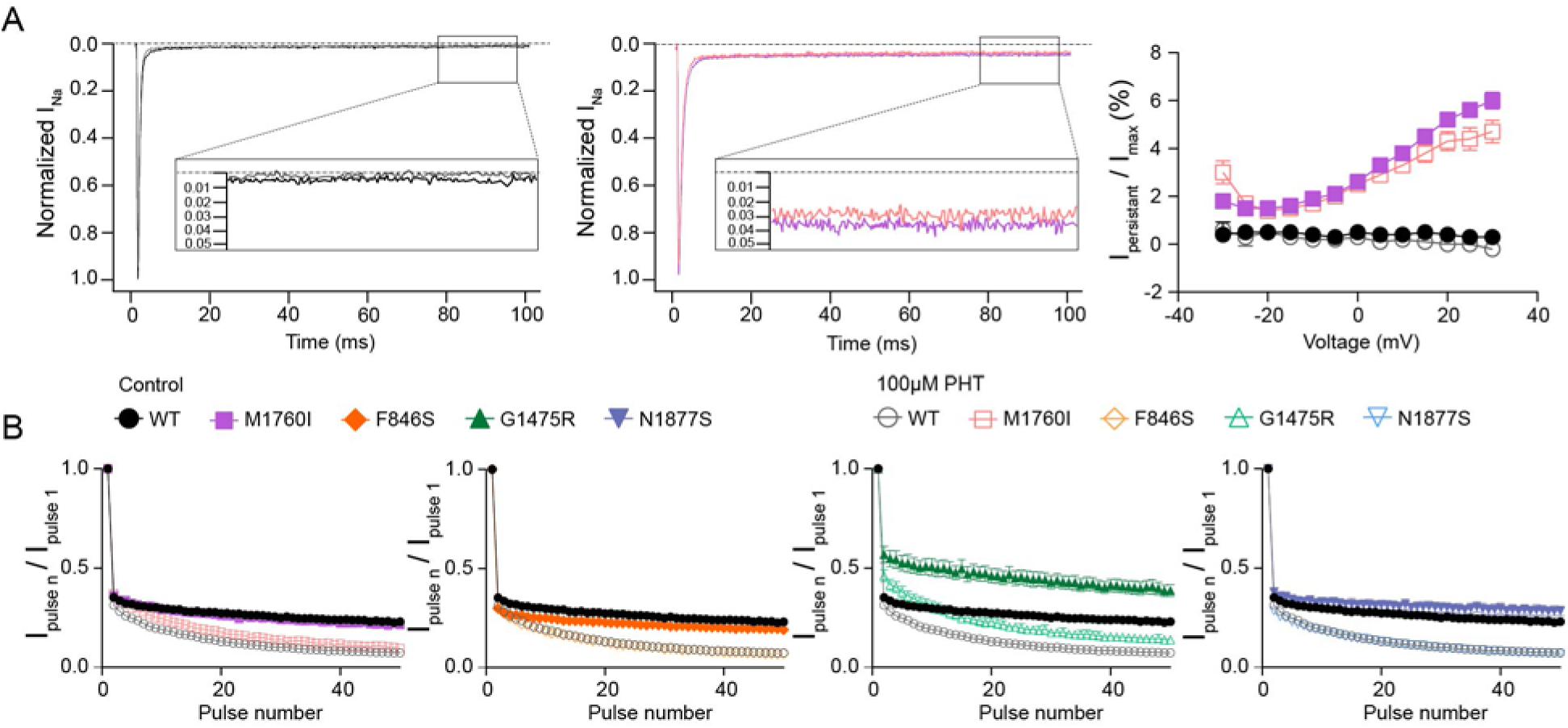
Effect of phenytoin on persistent current and use-dependence for WT and mutant Na_V_1.6 channels. (A) Left: Representative normalized traces of transient and persistent currents elicited by a step depolarization of +10 mV in the absence (black for WT, purple for M1760I) and presence (gray for WT, pink for M1760I) of 100 µM PHT. The insets show that the normalized persistent currents were measured between 75 ms and 95 ms. Right: The persistent currents normalized by the corresponding peak transient current amplitudes plotted against potential. (B) Use-dependence at 80Hz for WT and mutant Na_V_1.6 channels in the absence and presence of 100 µM PHT. All data are presented as means ± SEM.

**Supplementary Fig. 4:**
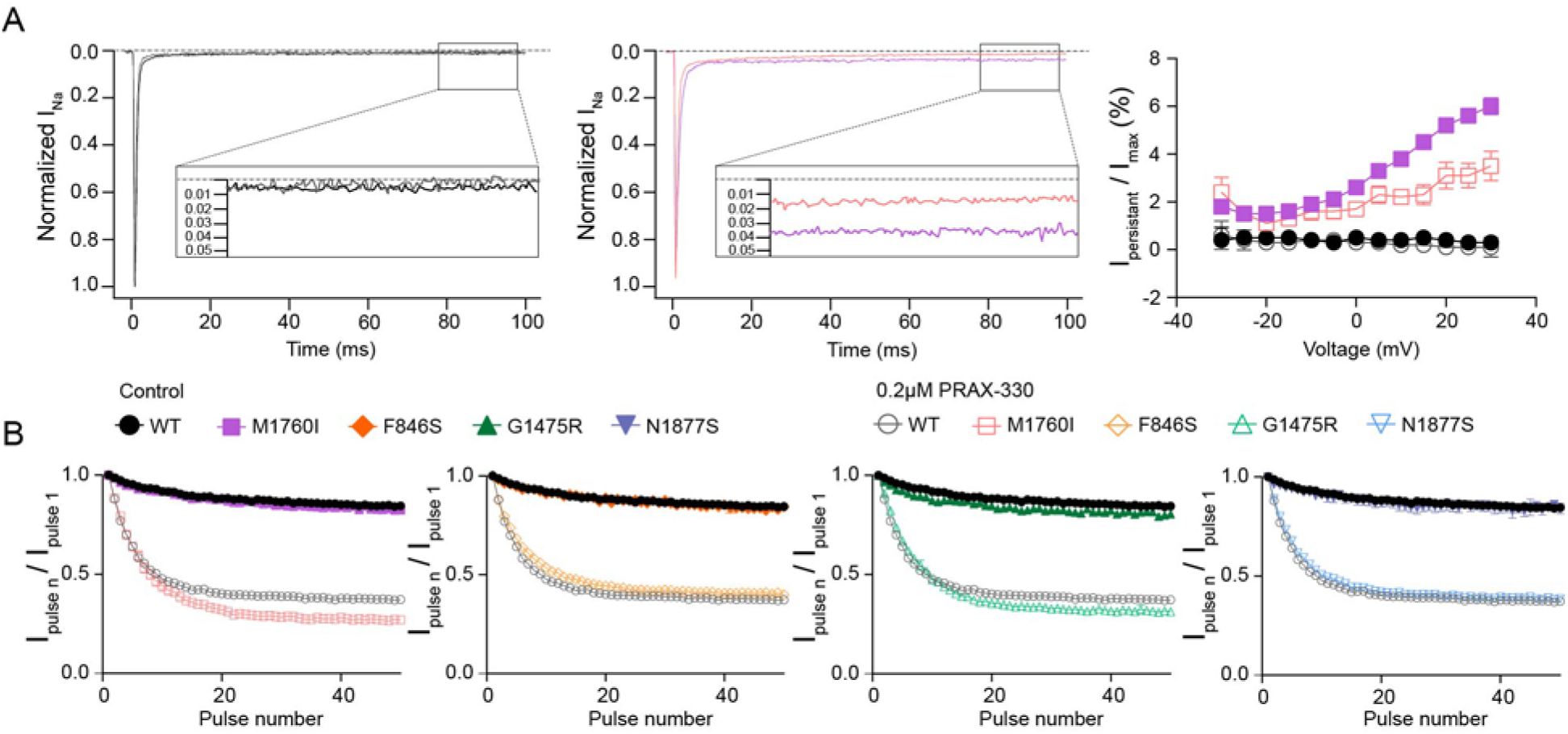
Effect of PRAX-330 on persistent current and use-dependent block for WT and mutant Na_V_1.6 channels. (A) Left: Representative normalized traces of transient and persistent currents elicited by a depolarization of +10 mV in the absence (black for WT, purple for M1760I) and presence (gray for WT, pink for M1760I) of 0.2 µM PRAX-330. The insets show that the normalized persistent currents were measured between 75 ms and 95 ms. Right: The persistent currents normalized by the corresponding peak transient current amplitudes plotted against potential. (B) Use-dependence at 20Hz for WT and mutant Na_V_1.6 channels in the absence and presence of 0.2 µM PRAX-330. All data are presented as means ± SEM.

## Notes

### Author Declarations

Ethics Committee of Region Zealand (SJ-91) and ethics committee of the Medical Faculty, University of Tuebingen (198/2010BO1) gave ethical approval for this work.

## References

1. Johannesen KM, Liu Y, Koko M, et al. Genotype-phenotype correlations in SCN8A-related disorders reveal prognostic and therapeutic implications. Brain. 2022;145(9):2991–3009. doi:10.1093/brain/awab321

2. Chung KM, Hack J, Andrews J, et al. Clinical severity is correlated with age at seizure onset and biophysical properties of recurrent gain of function variants associated with SCN8A-related epilepsy. Epilepsia. 2023;64(12):3365–3376. doi:10.1111/epi.17747

3. Millevert C, Weckhuysen S, ILAE Genetics Commission. ILAE Genetic Literacy Series: Self-limited familial epilepsy syndromes with onset in neonatal age and infancy. Epileptic Disord. 2023;25(4):445–453. doi:10.1002/epd2.20026

4. Anand G, Collett-White F, Orsini A, et al. Autosomal dominant SCN8A mutation with an unusually mild phenotype. Eur J Paediatr Neurol. 2016;20(5):761–765. doi:10.1016/j.ejpn.2016.04.015

5. Gardella E, Becker F, Møller RS, et al. Benign infantile seizures and paroxysmal dyskinesia caused by an SCN8A mutation. Ann Neurol. 2016;79(3):428–436. doi:10.1002/ana.24580

6. Liu Y, Schubert J, Sonnenberg L, et al. Neuronal mechanisms of mutations in SCN8A causing epilepsy or intellectual disability. Brain. 2019;142(2):376–390. doi:10.1093/brain/awy326

7. Johannesen KM, Gardella E, Encinas AC, et al. The spectrum of intermediate SCN8A-related epilepsy. Epilepsia. 2019;60(5):830–844. doi:10.1111/epi.14705

8. Gardella E, Marini C, Trivisano M, et al. The phenotype of SCN8A developmental and epileptic encephalopathy. Neurology. 2018;91(12):e1112–e1124. doi:10.1212/WNL.0000000000006199

9. Blanchard MG, Willemsen MH, Walker JB, et al. De novo gain-of-function and loss-of-function mutations of SCN8A in patients with intellectual disabilities and epilepsy. J Med Genet. 2015;52(5):330–337. doi:10.1136/jmedgenet-2014-102813

10. Wagnon JL, Barker BS, Hounshell JA, et al. Pathogenic mechanism of recurrent mutations of SCN8A in epileptic encephalopathy. Ann Clin Transl Neurol. 2016;3(2):114–123. doi:10.1002/acn3.276

11. Kwan P, Arzimanoglou A, Berg AT, et al. Definition of drug resistant epilepsy: consensus proposal by the ad hoc Task Force of the ILAE Commission on Therapeutic Strategies. Epilepsia. 2010;51(6):1069–1077. doi:10.1111/j.1528-1167.2009.02397.x

12. Ohba C, Kato M, Takahashi S, et al. Early onset epileptic encephalopathy caused by de novo SCN8A mutations. Epilepsia. 2014;55(7):994–1000. doi:10.1111/epi.12668

13. Qiao X, Sun G, Clare JJ, Werkman TR, Wadman WJ. Properties of human brain sodium channel α-subunits expressed in HEK293 cells and their modulation by carbamazepine, phenytoin and lamotrigine. Br J Pharmacol. 2014;171(4):1054–1067. doi:10.1111/bph.12534

14. Kuo CC. A common anticonvulsant binding site for phenytoin, carbamazepine, and lamotrigine in neuronal Na+ channels. Mol Pharmacol. 1998;54(4):712–721.

15. Kuo CC, Bean BP. Slow binding of phenytoin to inactivated sodium channels in rat hippocampal neurons. Mol Pharmacol. 1994;46(4):716–725.

16. Kuo CC, Chen RS, Lu L, Chen RC. Carbamazepine inhibition of neuronal Na+ currents: quantitative distinction from phenytoin and possible therapeutic implications. Mol Pharmacol. 1997;51(6):1077–1083. doi:10.1124/mol.51.6.1077

17. Bayraktar E, Liu Y, Sonnenberg L, et al. In vitro effects of eslicarbazepine (S-licarbazepine) as a potential precision therapy on SCN8A variants causing neuropsychiatric disorders. Br J Pharmacol. 2023;180(8):1038–1055. doi:10.1111/bph.15981

18. Baker EM, Thompson CH, Hawkins NA, et al. The novel sodium channel modulator GS-458967 (GS967) is an effective treatment in a mouse model of SCN8A encephalopathy. Epilepsia. 2018;59(6):1166–1176. doi:10.1111/epi.14196

19. Wengert ER, Saga AU, Panchal PS, Barker BS, Patel MK. Prax330 reduces persistent and resurgent sodium channel currents and neuronal hyperexcitability of subiculum neurons in a mouse model of SCN8A epileptic encephalopathy. Neuropharmacology. 2019;158:107699. doi:10.1016/j.neuropharm.2019.107699

20. Bunton-Stasyshyn RKA, Wagnon JL, Wengert ER, et al. Prominent role of forebrain excitatory neurons in SCN8A encephalopathy. Brain. 2019;142(2):362–375. doi:10.1093/brain/awy324

21. Estacion M, O’Brien JE, Conravey A, et al. A novel de novo mutation of SCN8A (Nav1.6) with enhanced channel activation in a child with epileptic encephalopathy. Neurobiol Dis. 2014;69:117–123. doi:10.1016/j.nbd.2014.05.017

22. Pan Y, Cummins TR. Distinct functional alterations in SCN8A epilepsy mutant channels. J Physiol. 2020;598(2):381–401. doi:10.1113/JP278952

23. Barker BS, Ottolini M, Wagnon JL, Hollander RM, Meisler MH, Patel MK. The SCN8A encephalopathy mutation p.Ile1327Val displays elevated sensitivity to the anticonvulsant phenytoin. Epilepsia. 2016;57(9):1458–1466. doi:10.1111/epi.13461

24. Veeramah KR, O’Brien JE, Meisler MH, et al. De novo pathogenic SCN8A mutation identified by whole-genome sequencing of a family quartet affected by infantile epileptic encephalopathy and SUDEP. Am J Hum Genet. 2012;90(3):502–510. doi:10.1016/j.ajhg.2012.01.006

25. Guo QB, Zhan L, Xu HY, Gao ZB, Zheng YM. SCN8A epileptic encephalopathy mutations display a gain-of-function phenotype and divergent sensitivity to antiepileptic drugs. Acta Pharmacol Sin. 2022;43(12):3139–3148. doi:10.1038/s41401-022-00955-x

26. Scheffer IE, Berkovic S, Capovilla G, et al. ILAE classification of the epilepsies: Position paper of the ILAE Commission for Classification and Terminology. Epilepsia. 2017;58(4):512–521. doi:10.1111/epi.13709

27. Beniczky S, Trinka E, Wirrell E, et al. Updated classification of epileptic seizures: Position paper of the International League Against Epilepsy. Epilepsia. 2025;66(6):1804–1823. doi:10.1111/epi.18338

28. Lyu H, Boßelmann CM, Johannesen KM, et al. Clinical and electrophysiological features of SCN8A variants causing episodic or chronic ataxia. EBioMedicine. 2023;98:104855. doi:10.1016/j.ebiom.2023.104855

29. Potet F, Vanoye CG, George AL. Use-Dependent Block of Human Cardiac Sodium Channels by GS967. Mol Pharmacol. 2016;90(1):52–60. doi:10.1124/mol.116.103358

30. Cutts A, Savoie H, Hammer MF, et al. Clinical characteristics and treatment experience of individuals with SCN8A developmental and epileptic encephalopathy (SCN8A-DEE): Findings from an online caregiver survey. Seizure. 2022;97:50–57. doi:10.1016/j.seizure.2022.03.008

31. Landrum MJ, Lee JM, Benson M, et al. ClinVar: improving access to variant interpretations and supporting evidence. Nucleic Acids Res. 2018;46(D1):D1062–D1067. doi:10.1093/nar/gkx1153

32. Menezes LFS, Sabiá Júnior EF, Tibery DV, Carneiro LDA, Schwartz EF. Epilepsy-Related Voltage-Gated Sodium Channelopathies: A Review. Front Pharmacol. 2020;11:1276. doi:10.3389/fphar.2020.01276

33. Liu Y, Koko M, Lerche H. A SCN8A variant associated with severe early onset epilepsy and developmental delay: Loss- or gain-of-function? Epilepsy Res. 2021;178:106824. doi:10.1016/j.eplepsyres.2021.106824

34. Belardinelli L, Liu G, Smith-Maxwell C, et al. A novel, potent, and selective inhibitor of cardiac late sodium current suppresses experimental arrhythmias. J Pharmacol Exp Ther. 2013;344(1):23–32. doi:10.1124/jpet.112.198887

35. Sicouri S, Belardinelli L, Antzelevitch C. Antiarrhythmic effects of the highly selective late sodium channel current blocker GS-458967. Heart Rhythm. 2013;10(7):1036–1043. doi:10.1016/j.hrthm.2013.03.023

36. Anderson LL, Thompson CH, Hawkins NA, et al. Antiepileptic activity of preferential inhibitors of persistent sodium current. Epilepsia. 2014;55(8):1274–1283. doi:10.1111/epi.12657

37. Auffenberg E, Hedrich UB, Barbieri R, et al. Hyperexcitable interneurons trigger cortical spreading depression in an Scn1a migraine model. J Clin Invest. 2021;131(21):e142202. doi:10.1172/JCI142202

38. Müller P, Draguhn A, Egorov AV. Persistent sodium currents in neurons: potential mechanisms and pharmacological blockers. Pflugers Arch. 2024;476(10):1445–1473. doi:10.1007/s00424-024-02980-7

39. Kahlig KM, Chapman M, Petrou S. PRAX-628: A Novel Sodium Channel Blocker with Greater Potency and Activity Dependence Compared to Standard of Care. In: 2023.

40. Potet F, Egecioglu DE, Burridge PW, George AL. GS-967 and Eleclazine Block Sodium Channels in Human Induced Pluripotent Stem Cell-Derived Cardiomyocytes. Mol Pharmacol. 2020;98(5):540–547. doi:10.1124/molpharm.120.000048

41. Kahlig KM, Scott L, Hatch RJ, et al. The novel persistent sodium current inhibitor PRAX-562 has potent anticonvulsant activity with improved protective index relative to standard of care sodium channel blockers. Epilepsia. 2022;63(3):697–708. doi:10.1111/epi.17149

42. Johnson JP, Focken T, Khakh K, et al. NBI-921352, a first-in-class, NaV1.6 selective, sodium channel inhibitor that prevents seizures in Scn8a gain-of-function mice, and wild-type mice and rats. Elife. 2022;11:e72468. doi:10.7554/eLife.72468

