## Supplementary information for "Differential effects of sodium channel blockers on *SCN8A* gain-of-function variants associated with drug-responsive or -resistant epilepsy"

**Supplementary materials and methods**

### Cell culture and transfection

ND7/23 cells (Sigma-Aldrich, RRID: CVCL_4259) were cultured in Dulbecco’s modified Eagle nutrient medium (Invitrogen) supplemented with 10% fetal calf serum (PAN-biothech) and 200mM L-glutamine (Biochrom) at 37 °C in a humidified atmosphere containing 5% CO_2_. Wild-type (WT) or mutant human Na_V_1.6 α-subunit cDNA (4 μg) together with 0.2 μg human β1- and 0.2 μg β2-subunits (pCLH-hβ1-EGFP and pCLH-hβ2-CD8) were transfected into ND7/23 cells following the standard protocol for Lipofectamine 2000 (Invitrogen). Electrophysiological recordings were performed 48 h after transfection and only from cells with TTX-resistant Na^+^ current that tested positive for both green fluorescence and anti-CD8 antibody coated microbeads.

Hippocampal primary neurons were obtained and transfected as previously described. Briefly, a mixture of 1 μg WT or mutant *SCN8A* α-subunit cDNA and 0.1 μg GFP plasmid under the CamKII-promotor were transfected into hippocampal neuronal cultures following the standard protocol of Optifect (Invitrogen). After 48 h, electrophysiological recordings were performed in fluorescence-positive neurons.

### Electrophysiology

Standard whole-cell patch clamp recordings were performed in transfected ND7/23 cells and hippocampal neuronal cultures at room temperature (RT, 20-25 °C) using an Axopatch 200B amplifier, a Digidata 1440A digitizer and Clampex 10.2 (Axon Instruments) data acquisition software. For recordings in ND7/23 cells, 500nM TTX was added into the bath solution to block endogenous Na^+^ current. Currents were filtered at 5 kHz and digitized at 20 kHz. Borosilicate glass pipettes had resistances of 1.5 - 2.5 MΩ when filled with the pipette solution (in mM): 10 NaCl, 1 EGTA, 10 HEPES, 140 CsF; pH 7.3 was adjusted to with CsOH, osmolarity was adjusted to 310 mOsm with mannitol. The bath solution contained (in mM): 140 NaCl, 3 KCl, 1 MgCl_2_, 1 CaCl_2_, 10 HEPES, 20 TEACl, 5 CsCl, 0.1 CdCl_2_; pH 7.3 was adjusted to with CsOH, osmolarity was adjusted to 320 mOsm with mannitol.

Recordings in transfected neurons were performed in the absence of TTX at RT. For current-clamp recordings, signals were filtered at 10 kHz and sampled at 100 kHz. Borosilicate glass pipettes had resistances of 3–5 MΩ when filled with the pipette solution (in mM): 5 KCl, 4 ATP-Mg, 10 phosphocreatine, 0.3 GTP-Na, 10 HEPES, 125 K-gluconate, 2 MgCl_2_ and 10 EGTA; pH was adjusted to 7.2 with KOH, osmolarity was adjusted to 290 mOsm/kg. The bath solution contained (in mM): 125 NaCl, 25 NaHCO_3_, 2.5 KCl, 1 MgCl_2_, 2 CaCl_2_, 1.25 NaH_2_PO_4_, 5 HEPES, 10 Glucose; pH was adjusted to 7.4 with NaOH, osmolarity was adjusted to 305 mOsm.

### Electrophysiological protocols and data analysis for recordings in ND7/23 cells

1. Current density, voltage-dependent steady-state activation and persistent current

Na^+^ currents were elicited by a series of 100 ms depolarizing voltage steps ranging from -80 mV to +65 mV in 5 mV increments from a holding potential of -100 mV. Current density was determined by normalizing peak Na^+^ currents to cell capacitance. The activation curve (conductance–voltage relationship) was derived from the current–voltage relationship obtained according to:

$$g\left( V \right)=\frac{I\left( V \right)}{V-V_{rev}}$$

where g is the conductance at test potential V, I the recorded peak Na^+^ current, and V_rev_ the observed Na^+^ reversal potential. A standard Boltzmann function was fit to the activation curves:

$$g\left( V \right)=\frac{g_{max}}{1+e[\frac{\left( V-V_{1/2} \right)}{k}]}$$

where g_max_ is the normalized peak conductance, V_1/2_ the voltage of half-maximal activation, and k the slope factor.

Persistent Na^+^ currents were determined at the end of depolarization pulses (75-95 ms) normalized to peak currents.

2. Voltage-dependent steady-state inactivation

Steady-state inactivation was determined using 2500-ms conditioning pulses ranging from -130 mV to -10 mV in 10 mV increments, followed by the test pulse to -10 mV at which the peak current reflected the percentage of non-inactivated channels. A standard Boltzmann function was fit to the inactivation curves:

$$I\left( V \right)=\frac{I_{max}}{1+e[\frac{\left( V-V_{1/2} \right)}{k}]}$$

where I is the recorded Na^+^ current amplitude at the conditioning potential V, I_max_ the maximal current amplitude, V_1/2_ the voltage of half-maximal inactivation, and k the slope factor.

3. Use-dependence

Use-dependence was examined using 50 depolarizing pulses to -20 mV at 20 Hz or 80 Hz. Peak currents of each pulse were normalized to the currents elicited by the first pulse to quantify cumulative Na^+^ current reduction.

4. Binding and unbinding kinetics

The protocols used to determine drug binding and unbinding kinetics were adapted from previously published studies^1^.

The voltage-step protocol used for assessing binding kinetics of drugs was shown in Supplementary Fig. 1A. The cells were first depolarized to -40 mV for variable durations ranging from 10 to 2500 ms, enabling the drug binding to the inactivated Na_V_ channels. This depolarization was followed by a brief hyperpolarizing step to −100 mV for 5 ms, allowing non-blocked channels to recover from fast inactivation. Subsequently, a test depolarizing pulse to -10 mV for 25 ms was applied to elicit Na^+^ currents (supplementary Fig. 1A), which were normalized and plotted against the duration of conditioning depolarization pulses (Supplementary Fig. 1B). The fraction of blocked current was determined by subtracting the normalized peak current obtained in the presence of the drug from the normalized current recorded in control solution (supplementary Fig. 1C). A first-order exponential function was fit to the fraction of blocked current plotted against the duration of the conditioning pulses (supplementary Fig. 1C). The reciprocal of the resulting time constant (τ_on_) represents the apparent binding rate, which increases linearly with drug concentration; the slope of this linear regression provides an estimate of the binding rate constant (supplementary Fig. 1D).

The voltage-step protocol used for assessing unbinding kinetics of drugs was shown in Supplementary Fig. 1E. The cells were first depolarized to -40 mV for 2 s to ensure a complete binding of the drug to the inactivated Na_V_ channels. This was followed by a hyperpolarizing step to -100 mV with different intervals (25-1000 ms for phenytoin, 250-4000 ms for PRAX-330), allowing the recovery from inactivation as well as the dissociation of drugs from inactivated channels. Because the recovery from inactivation is faster than the drug unbinding under these conditions, unbinding was the rate-limiting process. Finally, a test pulse to -10 mV was applied to assess channel availability (supplementary Fig. 1E). The fraction of blocked current was obtained by subtracting normalized currents recorded in the presence of the drug from normalized currents recorded in control solution. A first-order exponential function was fit to the relatively blocked current plotted against intervals and the resulting time constant was taken as the unbinding rate (τ_off_) of the drug (supplementary Fig. 1F). In contrast to the binding rate, the unbinding rate was independent of drug concentration revealed by similar unbinding rates of different drug concentrations.

### Electrophysiological protocols and data analysis for recordings in primary neuronal cultures

For experiments in transfected neurons, input resistance was determined by the slope of a linear regression fit to corresponding voltage responses plotted against a series of current injections (500 ms) ranging from -110 to -10 pA with 10 pA increments.

Action potential firing was examined using a series of current injections (800 ms) ranging from -50 to 300 pA with 25 pA increments. Action potentials were defined as events with peak voltages exceeding 0 mV. The threshold of an action potential was determined as the voltage at which the first derivative dV/dt reached 50 V/s^2^.

### Reference

1.	Qiao X, Sun G, Clare JJ, Werkman TR, Wadman WJ. Properties of human brain sodium channel α-subunits expressed in HEK293 cells and their modulation by carbamazepine, phenytoin and lamotrigine. *Br J Pharmacol*. 2014;171(4):1054-1067. doi:10.1111/bph.12534

2.	Kole MHP, Stuart GJ. Is action potential threshold lowest in the axon? *Nat Neurosci*. 2008;11(11):1253-1255. doi:10.1038/nn.2203

**Supplementary figures**

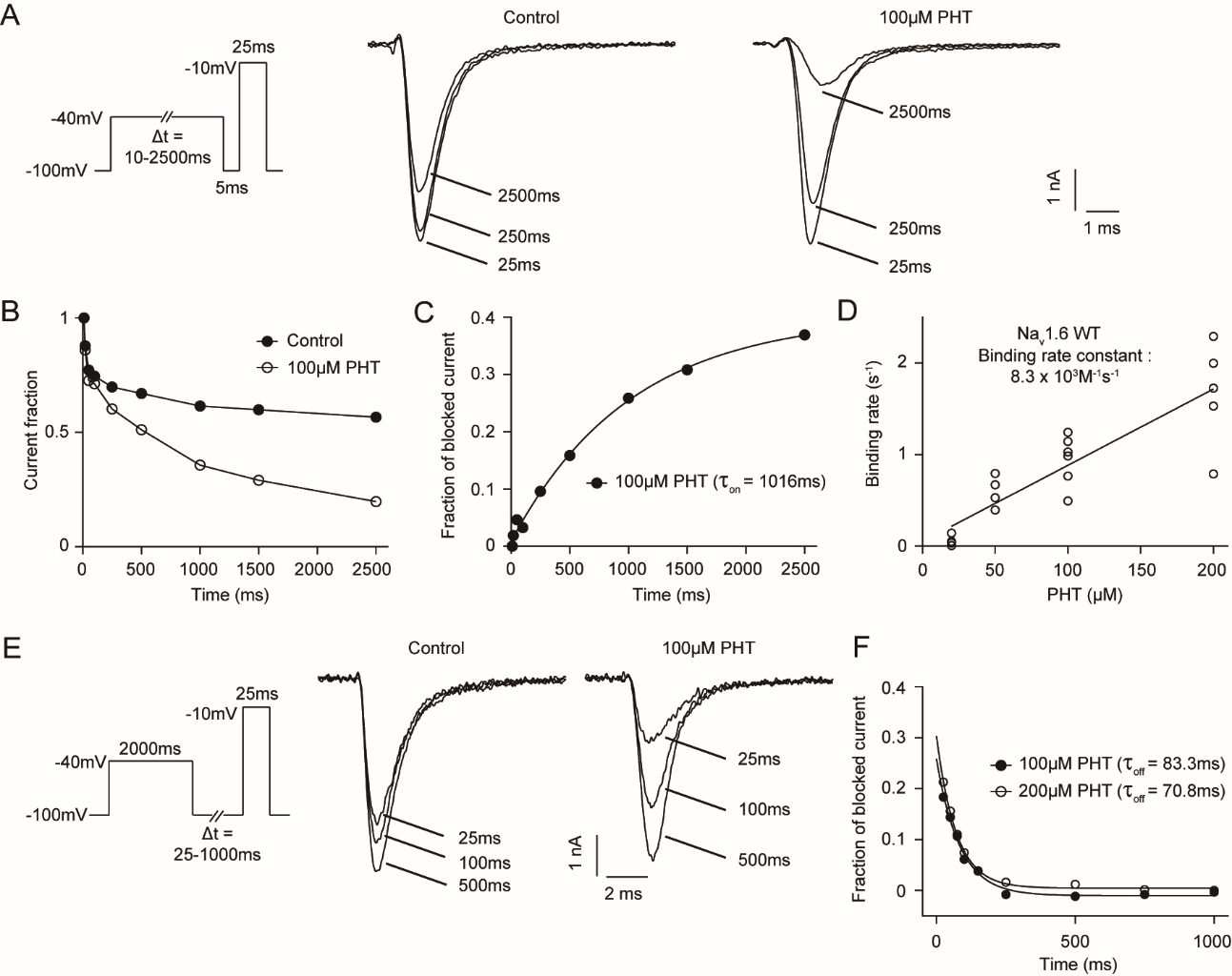

**Supplementary Figure 1: The binding and unbinding kinetics of phenytoin for Na_V_1.6 WT channel.** (A) Voltage-clamp protocol and representative traces of Na^+^ currents evoked by the test pulse to −10 mV following a 25, 250 or 2500 ms conditioning pulse to −40 mV and a short hyperpolarizing pulse for recovery from fast inactivation, both in the absence (control) and the presence of 100 μM PHT. (B) Normalized Na^+^ currents from a single cell in the absence and presence of 100 μM PHT plotted versus the pre-pulse duration time. (C) The relatively blocked Na^+^ current was derived from (B), which was determined by subtracting the normalized current recorded in the presence of PHT from the normalized current recorded in control solution. A first-order exponential function was fit to the data points, which were plotted against the pre-pulse duration, to determine the time constant (τ_on_) for development of block in the presence of 100 μM PHT. (D) Binding rates (1/τ_on_, s^−1^) in individual cells expressing WT Na_V_1.6 channels are plotted against the PHT concentrations. The slope of the linear regression provides an estimate of the binding rate constant. (E) Voltage-clamp protocol and representative traces of Na^+^ currents evoked by a 25 ms test pulse to −10 mV, following a 2000 ms conditioning pulse to -40 mV to allow full drug binding and 25, 100 or 500 ms hyperpolarizing pulses to −100 mV for recovery, both in the absence and presence of 100 μM PHT. (F) The relatively blocked current from a single cell was determined by subtracting the normalized current recorded in the presence of PHT from the normalized current recorded in control solution. A first-order exponential function was fit to the data points, which were plotted against the duration of the recovery pulses, to determine the time constant (τ_off_) of PHT unbinding. The similar time constants at 100 or 200 µM PHT showing that the unbinding rate is independent of the PHT concentration.

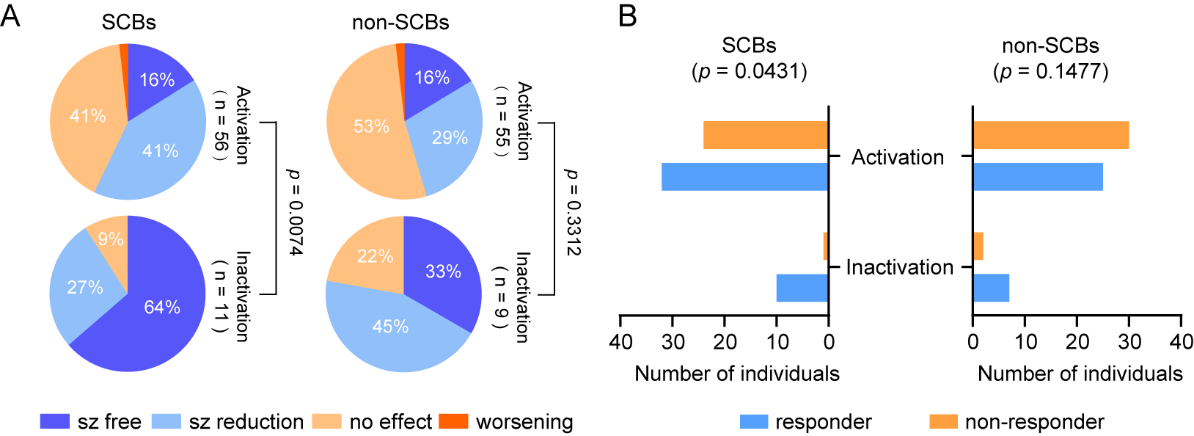

**Supplementary Fig. 2: Pharmacological responsiveness of individuals with severe DEE and ID.** (A) and (B) showed the treatment response to SCBs and non-SCBs in individuals with DEE and severe ID. P values derived from χ2 tests are provided under the pie chart or from Fisher´s exact tests are provided above the bar chart.

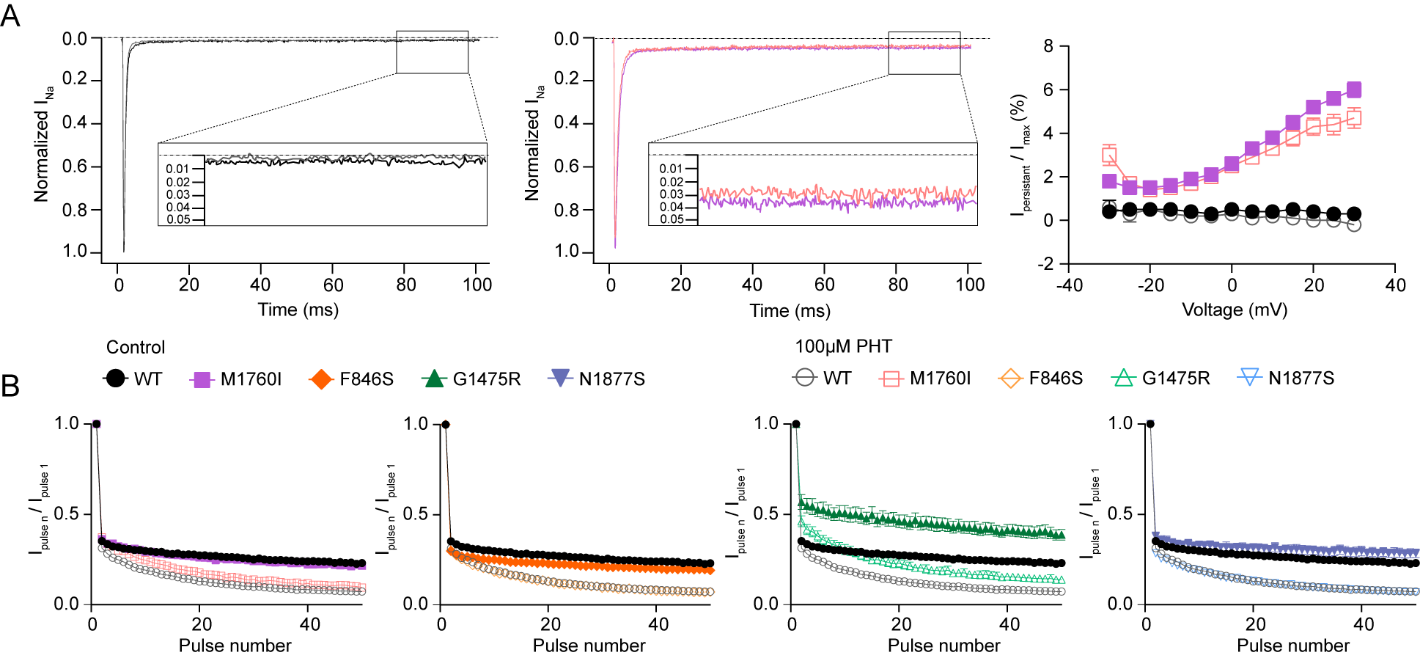

**Supplementary Fig. 3: Effect of phenytoin on persistent current and use-dependence for WT and mutant Na_V_1.6 channels.** (A) Left: Representative normalized traces of transient and persistent currents elicited by a step depolarization of +10 mV in the absence (black for WT, purple for M1760I) and presence (gray for WT, pink for M1760I) of 100 µM PHT. The insets show that the normalized persistent currents were measured between 75 ms and 95 ms. Right: The persistent currents normalized by the corresponding peak transient current amplitudes plotted against potential. (B) Use-dependence at 80Hz for WT and mutant Na_V_1.6 channels in the absence and presence of 100 µM PHT. All data are presented as means ± SEM.

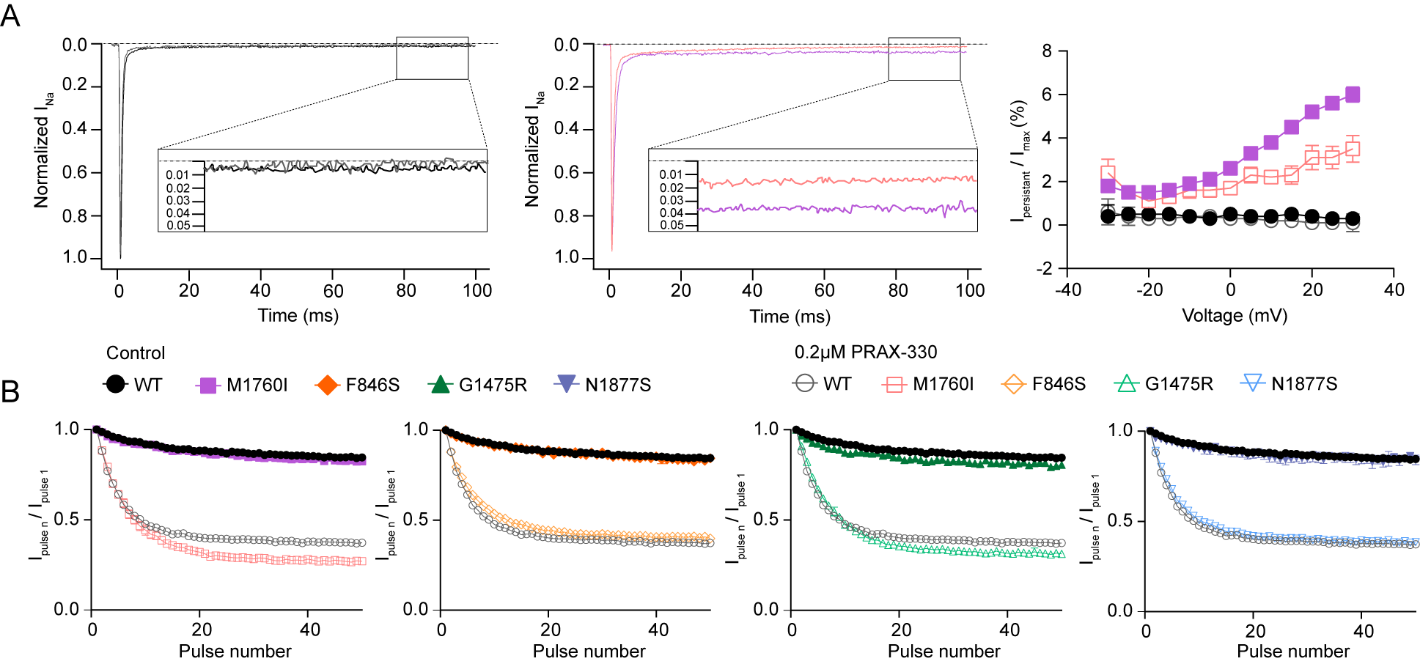

**Supplementary Fig. 4: Effect of PRAX-330 on persistent current and use-dependent block for WT and mutant Na_V_1.6 channels.** (A) Left: Representative normalized traces of transient and persistent currents elicited by a depolarization of +10 mV in the absence (black for WT, purple for M1760I) and presence (gray for WT, pink for M1760I) of 0.2 µM PRAX-330. The insets show that the normalized persistent currents were measured between 75 ms and 95 ms. Right: The persistent currents normalized by the corresponding peak transient current amplitudes plotted against potential. (B) Use-dependence at 20Hz for WT and mutant Na_V_1.6 channels in the absence and presence of 0.2 µM PRAX-330. All data are presented as means ± SEM.

**Supplementary tables**

**Supplementary Table 1.** Overview of the cohort of individuals carrying *SCN8A* gain-of-function variants

See excel file

**Supplementary Table 2.** Biophysical properties of *SCN8A* WT and mutant channels in ND7/23 cell in the absence or the presence of PHT

|  | Steady-state activation | | | Steady-state inactivation | | | Current density, pA/pF, (n) | Persistent current, % of peak current at +10 mV, (n) | Binding rate constant,  10^3^ M^-1^ s^-1^, (n) | Unbinding rate constant, s^-1^, (n) |
| --- | --- | --- | --- | --- | --- | --- | --- | --- | --- | --- |
|  | V ½, mV | k | n | V ½, mV | k | n |  |  |  |  |
| WT control | -20.8 ± 0.8 | -5.0 ± 0.3 | 18 | -69.2 ± 0.9 | 5.1 ± 0.2 | 15 | -278.6±25.6 (18) | 0.6 ± 0.1 (18) | 8.3 ± 1.2 (8) | 20.0 ± 2.7 (6) |
| WT PHT | -20.1 ± 1.4 | -4.7 ± 0.4 | 10 | -79.6 ± 0.9^####^ | 5.9 ± 0.2 | 9 | -241.5±32.6 (10) | 0.5 ± 0.1 (10) |  |  |
| M1760I control | -25.9 ± 1.4^****^ | -5.1 ± 0.3 | 16 | -69.5 ± 1.0 | 5.5 ± 0.2 | 10 | -317.3±41.9 (16) | 3.8 ± 0.2 (16) ^****^ | 17.2 ± 1.4^b^ (7) | 21.7 ± 3.5 (5) |
| M1760I PHT | -26.3 ± 1.0^****^ | -5.4 ± 0.3 | 12 | -80.1 ± 1.1^####^ | 6.5 ± 0.2 | 8 | -288.5±19.5 (12) | 3.3 ± 0.3 (12) ^****^ |  |  |
| F846S control | -30.0 ± 1.1^****^ | -5.7 ± 0.4 | 12 | -70.8 ± 0.7 | 5.9 ± 0.2 | 11 | -260.8 ± 31.4 (12) | 0.5 ± 0.2 (12) | 17.7 ± 1.8^b^ (7) | 16.6 ± 2.2 (6) |
| F846S PHT | -30.6 ± 1.7^****^ | -5.2 ± 0.4 | 10 | -79.7 ± 0.8^####^ | 6.1 ± 0.1 | 8 | -227.5±46.4 (10) | 0.5 ± 0.1 (10) |  |  |
| G1475R control | -19.1 ± 1.3 | -5.1 ± 0.4 | 16 | -62.1 ± 1.1^****^ | 5.5 ± 0.2 | 10 | -276.3±46.5 (16) | 0.6 ± 0.2 (16) | 14.8 ± 2.1 (6) | 21.1 ± 3.3 (5) |
| G1475R PHT | -19.3 ± 1.1 | -4.9 ± 0.4 | 10 | -72.0 ± 0.9^****,^  ^####^ | 5.6 ± 0.2 | 11 | -241.7±45.3 (10) | 0.6 ± 0.2 (10) |  |  |
| N1877S control | -22.5 ± 1.7 | -5.2 ± 0.4 | 12 | -66.5 ± 1.5^***^ | 5.5 ± 0.5 | 10 | -245.9±46.3 (12) | 0.4 ± 0.1 (12) | 11.2 ± 1.3 (6) | 20.2 ± 2.8 (6) |
| N1877S PHT | -21.9 ± 1.5 | -4.7 ± 0.3 | 10 | -77.0 ± 1.0^*,####^ | 5.8 ± 0.3 | 9 | -221.0±34.5 (10) | 0.4 ± 0.1 (12) |  |  |

Data are presented as means ± SEM; n = number of recorded cells; For comparisons among genotypes (WT vs. mutant), significance is indicated as ^*^*P* < 0.05, ^**^*P* < 0.01, ^***^*P* < 0.001, ^****^*P* < 0.0001; For comparisons before vs. after drug treatment within the same genotype, significance is indicated as ^#^*P* < 0.05, ^##^*P* < 0.01, ^###^*P* < 0.001, ^####^*P* < 0.0001; For comparisons of binding rate constants (WT vs. mutant), significance is indicated as ^a^*P* < 0.05, ^b^*P* < 0.01, ^c^*P* < 0.001, ^d^*P* < 0.0001.

**Supplementary Table 3.** Neuronal properties in excitatory hippocampal neurons in the absence or the presence of PHT

|  | Input resistance, MΩ | Resting membrane potential (RMP), mV | Threshold, mV | I/O area under the curve, nA | n |
| --- | --- | --- | --- | --- | --- |
| WT control | 271.6 ± 12.4 | -66.1 ± 0.8 | -41.8 ± 0.8 | 4.03 ± 0.26 | 16 |
| WT PHT | 266.5 ± 20.1 | -66.4 ± 0.8 | -41.9 ± 0.8 | 2.93 ± 0.26^#^ | 14 |
| M1760I control | 236.0 ± 13.1 | -66.4 ± 1.1 | -45.5 ± 0.9^***^ | 4.37 ± 0.20 | 16 |
| M1760I PHT | 232.2 ± 15.4 | -68.5 ± 1.0 | -44.9 ± 1.0 | 4.45 ± 0.23 | 14 |
| F846S control | 224.8 ± 16.1 | -69.2 ± 1.1 | -46.8 ± 0.8^****^ | 4.49 ± 0.25 | 17 |
| F846S PHT | 223.1 ± 13.8 | -66.1 ± 1.0 | -46.3 ± 1.0 | 4.12 ± 0.32 | 13 |
| G1475R control | 268.4 ± 15.7 | -66.4 ± 1.0 | -41.9 ± 0.6 | 4.28 ± 0.37 | 16 |
| G1475R PHT | 262.3 ± 10.7 | -66.7 ± 0.8 | -40.1 ± 0.6 | 2.75 ± 0.27^###^ | 17 |
| N1877S control | 224.3 ± 14.5 | -67.5 ± 1.4 | -41.2 ± 0.8 | 4.37 ± 0.37 | 13 |
| N1877S PHT | 235.1 ± 14.7 | -68.4 ± 0.9 | -40.6 ± 0.6 | 2.90 ± 0.18^##^ | 12 |

I/O = input-output curve. Data are presented as means ± SEM; n = number of recorded cells; For comparisons among genotypes (WT vs. mutant), significance is indicated as ^*^*P* < 0.05, ^**^ *P*< 0.01, ^***^*P* < 0.001, ^****^*P* < 0.0001; For comparisons before vs. after drug treatment within the same genotype, significance is indicated as ^#^*P* < 0.05, ^##^*P* < 0.01, ^###^*P* < 0.001, ^####^*P* < 0.0001.

**Supplementary Table 4.** Biophysical properties of *SCN8A* WT and mutant channels in ND7/23 cell in the absence or the presence of PRAX330

|  | Steady-state activation (10s) | | | Steady-state activation (2s) | | | Steady-state inactivation | | | Current density, pA/pF, (n) | Persistent current, % of peak current at +10 mV, (n) | Binding rate constant,  10^6^M^-1^s^-1^, (n) | Unbinding rate constant, s^-1^, (n) |
| --- | --- | --- | --- | --- | --- | --- | --- | --- | --- | --- | --- | --- | --- |
|  | V ½, mV | k | n | V ½, mV | k | n | V ½, mV | k | n |  |  |  |  |
| WT control | -18.6 ± 0.9 | -5.3 ± 0.1 | 11 | -20.8 ± 0.8 | -5.0 ± 0.3 | 18 | -69.2 ± 0.9 | 5.1 ± 0.2 | 15 | -278.6 ± 25.6 (18) | 0.6 ± 0.1 (18) | 24.1 ± 1.9 (7) | 1.24 ± 0.10 (7) |
| WT PRAX330 | -19.0 ± 1.4 | -5.3 ± 0.2 | 7 | -17.2 ± 1.6 | -6.4 ± 0.3^####^ | 7 | -78.0 ± 1.6^####^ | 5.3 ± 0.2 | 8 | -241.5 ± 32.6 (10) | 0.6 ± 0.3 (10) |  |  |
| M1760I control | -26.0 ± 1.1  ^****^ | -5.7 ± 0.3 | 8 | -25.9 ± 1.4  ^****^ | -5.1 ± 0.3 | 16 | -69.5 ± 1.0 | 5.5 ± 0.2 | 10 | -317.3 ± 41.9 (10) | 3.8 ± 0.2 (16)  ^****^ | 37.2 ± 2.5^d^ (6) | 1.25 ± 0.11 (6) |
| M1760I PRAX330 | -25.2 ± 1.  2^****^ | -5.4 ± 0.3 | 8 | -23.6 ± 1.1  ^****^ | -6.2 ± 0.6^##^ | 11 | -77.2 ± 0.9^####^ | 6.1 ± 0.2 | 7 | -280.1 ± 22.2 (8) | 2.2 ± 0.3 (10)  ^****, ####^ |  |  |
| F846S control | -28.6 ± 1.4  ^****^ | -6.0 ± 0.4 | 10 | -30.0 ± 1.1  ^****^ | -5.7 ± 0.4 | 12 | -70.8 ± 0.7 | 5.9 ± 0.2 | 11 | -260.8 ± 31.4 (9) | 0.5 ± 0.2 (12) | 27.8 ± 2.5 (6) | 1.46 ± 0.21 (6) |
| F846S PRAX330 | -28.0 ± 1.5  ^****^ | -5.9 ± 0.4 | 9 | -27.0 ± 1.5  ^****^ | -6.4 ± 0.5^###^ | 9 | -79.7 ± 0.7^####^ | 6.1 ± 0.2 | 10 | -224.5 ± 22.8 (9) | 0.5 ± 0.3 (9) |  |  |
| G1475R control | -19.3 ± 1.6 | -5.0 ± 0.2 | 11 | -19.1 ± 1.3 | -5.1 ± 0.4 | 16 | -62.1 ± 1.1^****^ | 5.5 ± 0.2 | 10 | -276.3 ± 46.5 (11) | 0.6 ± 0.2 (16) | 24.2 ± 2.4 (6) | 1.41 ± 0.15 (5) |
| G1475R PRAX330 | -17.9 ± 1.2 | -4.9 ± 0.4 | 10 | -16.0 ± 1.4 | -6.1 ± 0.4^###^ | 10 | -69.6 ± 1.3  ^****,####^ | 5.6 ± 0.3 | 9 | -245.8 ± 31.8 (10) | 0.6 ± 0.2 (10) |  |  |
| N1877S control | -20.7 ± 1.5 | -5.0 ± 0.5 | 9 | -22.5 ± 1.7 | -5.2 ± 0.4 | 12 | -66.5 ± 1.5^***^ | 5.5 ± 0.5 | 10 | -245.9 ± 46.3 (7) | 0.4 ± 0.1 (12) | 26.1 ± 1.5 (6) | 1.56 ± 0.11 (6) |
| N1877S PRAX330 | -21.9 ± 1.5 | -4.7 ± 0.3 | 9 | -18.2 ± 1.6 | -6.5 ± 0.5  ^####^ | 6 | -74.0 ± 1.0^***,^  ^####^ | 5.8 ± 0.3 | 6 | -206.4 ± 44.5 (9) | 0.5 ± 0.1 (9) |  |  |

Data are presented as means ± SEM; n = number of recorded cells; For comparisons among genotypes (WT vs. mutant), significance is indicated as ^*^*P* < 0.05, ^**^*P* < 0.01, ^***^*P* < 0.001, ^****^*P* < 0.0001; For comparisons before vs. after drug treatment within the same genotype, significance is indicated as ^#^*P* < 0.05, ^##^*P* < 0.01, ^###^*P* < 0.001, ^####^*P* < 0.0001; For comparisons of binding rate constants (WT vs. mutant), significance is indicated as ^a^*P* < 0.05, ^b^*P* < 0.01, ^c^*P* < 0.001, ^d^*P* < 0.0001.

**Supplementary Table 5.** Neuronal properties in excitatory hippocampal neurons in the absence or the presence of PRAX330

|  | Input resistance, MΩ | Resting membrane potential (RMP), mV | Threshold, mV | I/O area under the curve, nA | n |
| --- | --- | --- | --- | --- | --- |
| WT control | 271.6 ± 12.4 | -66.1 ± 0.8 | -41.8 ± 0.8 | 4.03 ± 0.26 | 16 |
| WT PRAX330 | 243.2 ± 12.9 | -67.7 ± 1.1 | -39.3 ± 0.7 | 1.70 ± 0.16^####^ | 14 |
| M1760I control | 236.0 ± 13.1 | -64.4 ± 1.1 | -45.5 ± 0.9^***^ | 4.37 ± 0.20 | 16 |
| M1760I PRAX330 | 240.4 ± 20.6 | -66.1 ± 0.8 | -40.1 ± 0.6^####^ | 1.50 ± 0.24^####^ | 10 |
| F846S control | 224.8 ± 16.1 | -69.2 ± 1.1 | -46.8 ± 0.8^****^ | 4.49 ± 0.25 | 17 |
| F846S PRAX330 | 221.5 ± 16.8 | -68.0 ± 1.0 | -45.6 ± 0.9^****^ | 1.81 ± 0.23^####^ | 13 |
| G1475R control | 268.4 ± 15.7 | -66.4 ± 1.0 | -41.9 ± 0.6 | 4.28 ± 0.37 | 16 |
| G1475R PRAX330 | 233.1 ± 17.9 | -64.8 ± 0.6 | -40.6 ± 1.0 | 1.56 ± 0.24^####^ | 12 |
| N1877S control | 224.3 ± 14.5 | -67.5 ± 1.4 | -41.2 ± 0.8 | 4.37 ± 0.37 | 13 |
| N1877S PRAX330 | 240.0 ± 10.8 | -66.4 ± 0.8 | -40.9 ± 0.6 | 1.91 ± 0.23^####^ | 12 |

I/O = input-output curve. Data are presented as means ± SEM; n = number of recorded cells; For comparisons among genotypes (mutant vs. WT), significance is indicated as ^*^*P* < 0.05, ^**^*P* < 0.01, ^***^*P* < 0.001, ^****^*P* < 0.0001; For comparisons before vs. after drug treatment within the same genotype, significance is indicated as ^#^*P* < 0.05, ^##^*P* < 0.01, ^###^*P* < 0.001, ^####^*P* < 0.0001.
